# The COPI coatomer regulates several steps of HDL metabolism

**DOI:** 10.1101/2025.08.21.25332476

**Authors:** Grigorios Panteloglou, Paolo Zanoni, Christopher S. Law, Brian Woods, Alaa Othman, Mustafa Yalcinkaya, Simon F. Norrelykke, Andrzej J. Rzepiela, Szymon Stoma, Michael Stebler, Anja Kerksiek, Michele Visentin, Marieke Smit, Sofia Kakava, Anton Potapenko, Eveline Schlumpf, Silvija Radosavljevic, Marta Futema, Nawar Dalila, Anne Tybjaerg-Hansen, Steve E. Humphries, Jan Albert Kuivenhoven, Bart van de Sluis, Dieter Lütjohann, Roger Meier, Jérôme Robert, Janet Chou, Raif S. Geha, Anthony K. Shum, Lucia Rohrer, Arnold von Eckardstein

## Abstract

**Background:** Reverse cholesterol transport by high density lipoproteins (HDL) is considered as an anti-atherogenic metabolic pathway. Hepatocytes determine the efficacy of this pathway by the production of apolipoprotein A-I (apoA-I) and its lipidation by ATP binding cassette transporter A1 (ABCA1), selective uptake of cholesterol via scavenger receptor BI (SR-BI) and uptake of entire HDL particles. The molecular determinants of the latter step are not well understood.

**Methods:** We performed a genome-wide RNA interference screen for genes limiting the uptake of fluorescent HDL into Huh-7 hepatocarcinoma cells. Top hit genes were validated by targeted *in vitro* experiments and the analysis of associations between their variants and HDL cholesterol levels in the databases of the Global Lipids Genetics Consortium and UK Biobank as well as inborn errors of metabolism and their respective mouse models.

**Results:** The knockdown of 128 genes significantly inhibited HDL uptake. Six of them encode for components of the COPI coatomer, namely *COPA, COPB1, COPB2, COPG1, ARCN1,* and *COPZ1*. Knocking down any of them decreased the uptake of both fluorescently labeled proteins and lipids of HDL, the cell surface abundance of SR-BI as well as *APOA1* expression and apoA-I secretion but increased the cell surface abundance of ABCA1 as well as cholesterol efflux. Single nucleotide polymorphisms of *COPB1, ARCN1,* and *COPZ2* were associated with significantly higher HDL-cholesterol (HDL-C) levels in the population while rare COPA and COPG1 variants causing immunopathies in humans and mice were associated with low levels of HDL cholesterol.

**Conclusions:** In hepatocytes, the COPI coatomer regulates HDL holoparticle uptake, selective lipid uptake, apoA-I secretion, and cholesterol efflux, and thereby, it influences plasma levels of HDL-C.

**Highlights:**

- By genome-wide RNA interference screening and replication experiments we found six components of the COPI coatomer, namely *COPA, COPB1, COPB2, COPG1, ARCN1,* and *COPZ1* to limit the uptake of HDL holoparticles into Huh-7 hepatocarcinoma cells, possibly by a mechanism that involves ATP binding cassette transporter ABCA1
- Loss of any of expression of any of these six COPI genes decreases HDL lipid uptake by interfering with the glycosylation and cell surface abundance of scavenger receptor SR-BI
- Loss of any of expression of any of these six COPI genes decreases HDL secretion from Huh7 cells, possibly by decreasing gene expression of APOA1 and ABCA1, but despite increasing cell surface abundance of ABCA1 and ABCA1-mediated cholesterol efflux
- Single nucleotide polymorphisms of *ARCN1 are* associated with significantly higher HDL-cholesterol (HDL-C) levels in the population while rare COPA and COPG1 variants causing immunopathies in humans and mice were associated with rather low levels of HDL cholesterol.

## Introduction

Low plasma levels of HDL cholesterol are associated with increased risk of atherosclerotic cardiovascular disease (ASCVD), but unsuccessful drug developments and negative findings of Mendelian Randomization studies have led to doubts about the causal role of HDL-cholesterol in ASCVD^1^. Nevertheless, the mediation of reverse cholesterol transport by HDL is still considered as an important anti-atherogenic metabolic pathway. The first steps in this process are well understood, namely the biogenesis of HDL by secretion of lipid-free apoA-I, its lipidation by ATP binding cassette transporters ABCA1 and ABCG1, and the subsequent esterification of cholesterol by lecithin:cholesterol acyltransferase (LCAT) as well as the HDL-mediated cholesterol efflux from macrophage foam cells, ^1^. However, the mechanism of the final steps of reverse cholesterol transport, namely the hepatic uptake of HDL, is only partially resolved. Cholesteryl esters of HDL are indirectly delivered to the liver by cholesteryl ester transfer protein (CETP)-mediated transfer to apoB-containing lipoproteins that are ultimately taken up by the low density lipoprotein (LDL) receptor (LDL-R)^1,2^. The direct delivery involves both selective uptake of lipids independently of HDL’s protein moiety by SR-BI, and HDL-holoparticle uptake by a yet little understood mechanism^2^. The latter appears to involve high affinity binding of apoA-I, i.e. the main protein constituent of HDL, by ectopic beta-ATPase and thereby generation of ADP, which in turn stimulates the purinergic P2Y13 receptor to signal to a yet unidentified lower affinity HDL binding site, distinct from SR-BI, that mediates the endocytosis of HDL particles^2,3^.

To identify genes that limit the endocytosis of HDL by the liver, we performed a genome wide siRNA screening in Huh-7 hepatocarcinoma cells. The uptake of HDL was most significantly inhibited by interference with the *COPA, COPB1, COPB2, COPG1, ARCN1,* and *COPZ1* genes which encode for indispensable subunits of the COPI coatomer, namely α-COP, β-COP, β’-COP, γ-COP, δ-COP and ζ-COP, respectively^4^. β-COP, δ-COP, γ-COP and ζ-COP form the F-subcomplex, while α-COP, β’-COP, and the thermosensitive and accessory ε-COP form the B-subcomplex (figure 1A). Together the B- and F-subcomplexes encase vesicles mediating the transport of proteins either from the Golgi apparatus to the endoplasmic reticulum or between the Golgi cisternae, and thereby play an important role in the processing and quality control of proteins^4^. In addition, COPI contributes to the maturation of early endosomes, to lysosomal trafficking and autophagy^5,6^. Rare variants in *COPA* and *COPG1* underlie inherited autoinflammatory and immunodeficiency diseases, respectively^7–9^.

**Figure 1.**
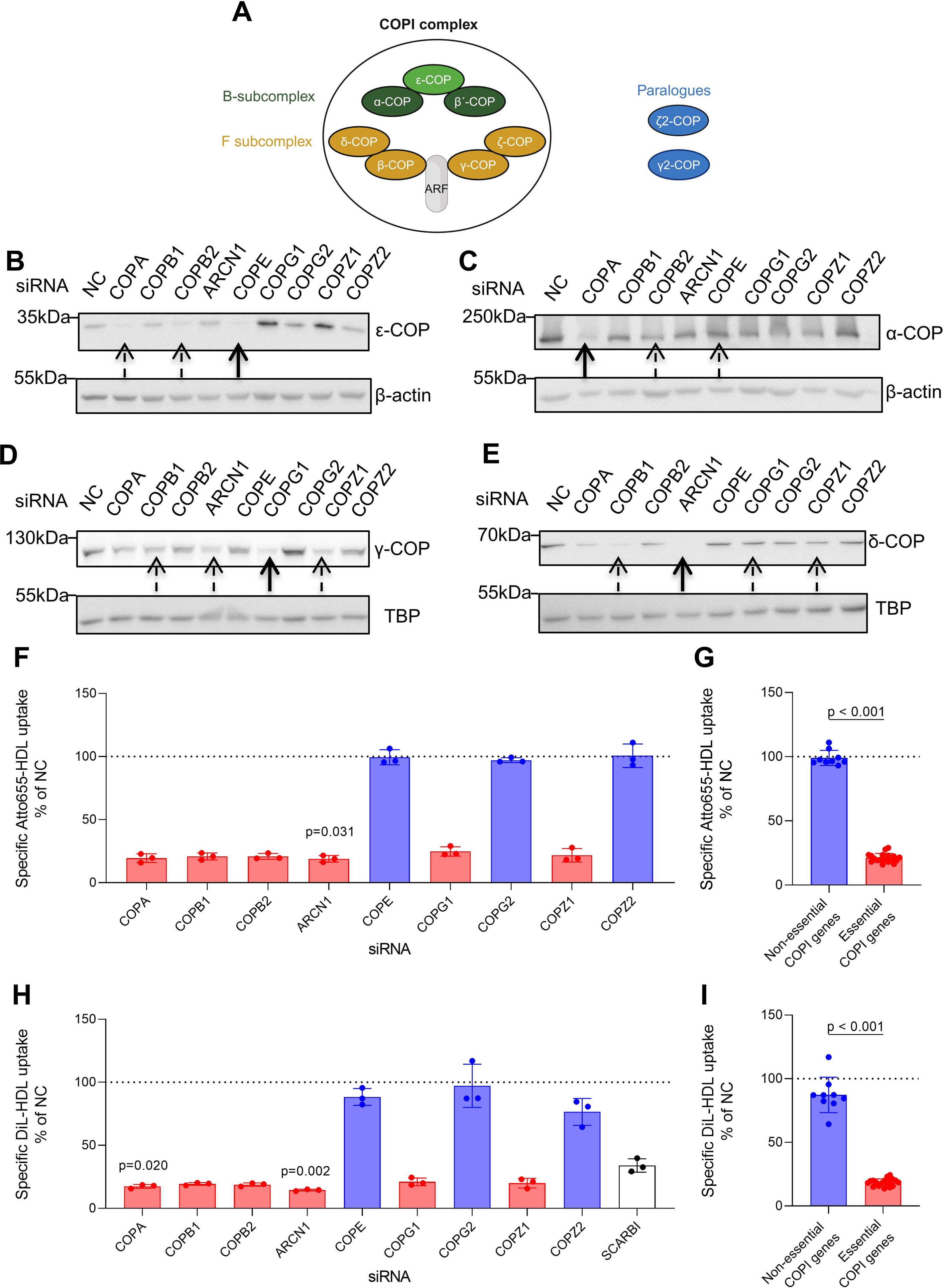
Silencing of COPI genes and its effect on the uptake of HDL into Huh-7 hepatocarcinoma cells. **A** Illustration depicting the formation of COPI complex as a heterodimer of sub complexes B (comprised of α-, β,- and ε-COP) and F (comprised of β-, γ-, δ-, and ζ-COP). γ2- and ζ2-COP, paralogues of γ- and ζ-COP respectively, are also shown. **B-E.** Huh-7 cells transfected with the indicated siRNAs were collected 72 hours post transfection. Lysates were used for Wester Blotting of ε-COP (**B**), α-COP (**C**), γ-COP (**D**) or δ-COP (**E**). β-actin (**B**,**C**) or TATA binding protein (TBP) (**D, E**) as the loading control. The Western Blots are representatives of two (**B**, **C**) or four (**E**) independent experiments, while the Western Blotting experiment in **D** was carried out only once. Expression of the other COPI proteins were not investigated because of lack of available antibodies. Bold arrows indicate the COPI protein directly targeted by the siRNA while hatched arrows indicate COPI proteins residing in the same B-subcomplex (**B, C**) or F-subcomplex (**D, E**) of the coatomer. **F-I.** 72 hours after transfection with the indicated siRNAs, Huh-7 cells were incubated with 20 μg/mL Atto655-HDL (**F**, **G**) or Dil-HDL (**H**, **I**) for 3 hours in the presence or absence of 100-fold excess unlabeled HDL and were then collected for measurement with flow cytometry. The specific cell association was calculated as the difference between the two conditions. The data were normalized to the non-coding control (NC). *SCARBI* (**H**) was included as the positive control. **F** and **H** show means ± SD of 3 independent experiments for each condition separately. Statistical analysis was performed using Kruskal-Wallis test coupled with Dunn’s test for multiple comparisons between the NC and each targeting siRNA. **G** and **I** compare the summarized data on the non-dispensable COPI subunits (red bars) with the data on the dispensable or paralogous COPI genes (blue bars) by using two-tailed Mann-Whitney test (**G**) or two-tailed unpaired t-test (**I**). Only statistically significant differences (p<0.05) are indicated.

Since the role of the COPI coatomer in lipoprotein metabolism is little understood, we investigated the cellular mechanisms underlying disturbed HDL uptake upon loss of COPI function, as well as the impact of genetic variants in COPI genes on plasma HDL-C concentrations in humans and mice.

## Methods and Data Availability

All data and methods supporting the findings of this study are available in the Online Data Supplement or from the corresponding authors on reasonable request.

## Results

### Genome wide RNAi screen identifies several COPI components as limiting factors of HDL endocytosis

We performed a genome-wide siRNA screen in Huh-7 hepatocarcinoma cells for genes that limit the uptake of HDL containing a fluorescently labeled protein moiety. Since no receptor is known to mediate the endocytosis of HDL holoparticle into hepatocytes^2^ and our exploratory pilot experiments also excluded a limiting effect of RNA interference against *SCARB1,* which was shown to facilitate HDL holoparticle uptake by endothelial cells but not hepatocytes^10^, we could not include any condition to control for specificity. As in our LDL screen^11^, we applied Redundant siRNA Activity (RSA) analysis to data from the best performing assay feature, namely median cytoplasm intensity, for the identification of hit genes. Z’-factor values^11^ for median cytoplasm intensity in each assay plate for the background without fluorescent HDL (median 0.79, interquartile range [IQR] 0.68 to 0.90) indicated excellent signal-to-noise ratio (supplemental figure S1).

At the RSA p value cut off of p < 0.001, silencing of 128 and 47 genes decreased and increased HDL uptake, respectively (table 1 and supplemental table S1). The three top hits for decreased HDL uptake were *COPA, ARCN1,* and *COPZ1*, which encode for three of the seven canonical components of the COPI coatomer complex. Silencing of *COPB1* (rank 8), *COPB2* (rank 11), and *COPG1* (rank 23) also significantly decreased the uptake of HDL in Huh-7 cells (table 1). Silencing of COPE, which encodes for the thermosensitive and accessory ε-COP subunit of the COPI coatomer’s B-subcomplex^12,13^, also resulted in lower HDL uptake, however, without passing the RSA p-value cut off (supplemental table S1). The knockdowns of *COPG2* and *COPZ*2 which encode for paralogues of γ-COP and ζ-COP, respectively, but are not part of the COPI coatomer^14,15^ did not reduce HDL uptake (supplemental table S1). Of note, *SCARB1* belonged to the genes whose knockdown increased the uptake of HDL in Huh-7 cells (table 1).

**Table 1.**
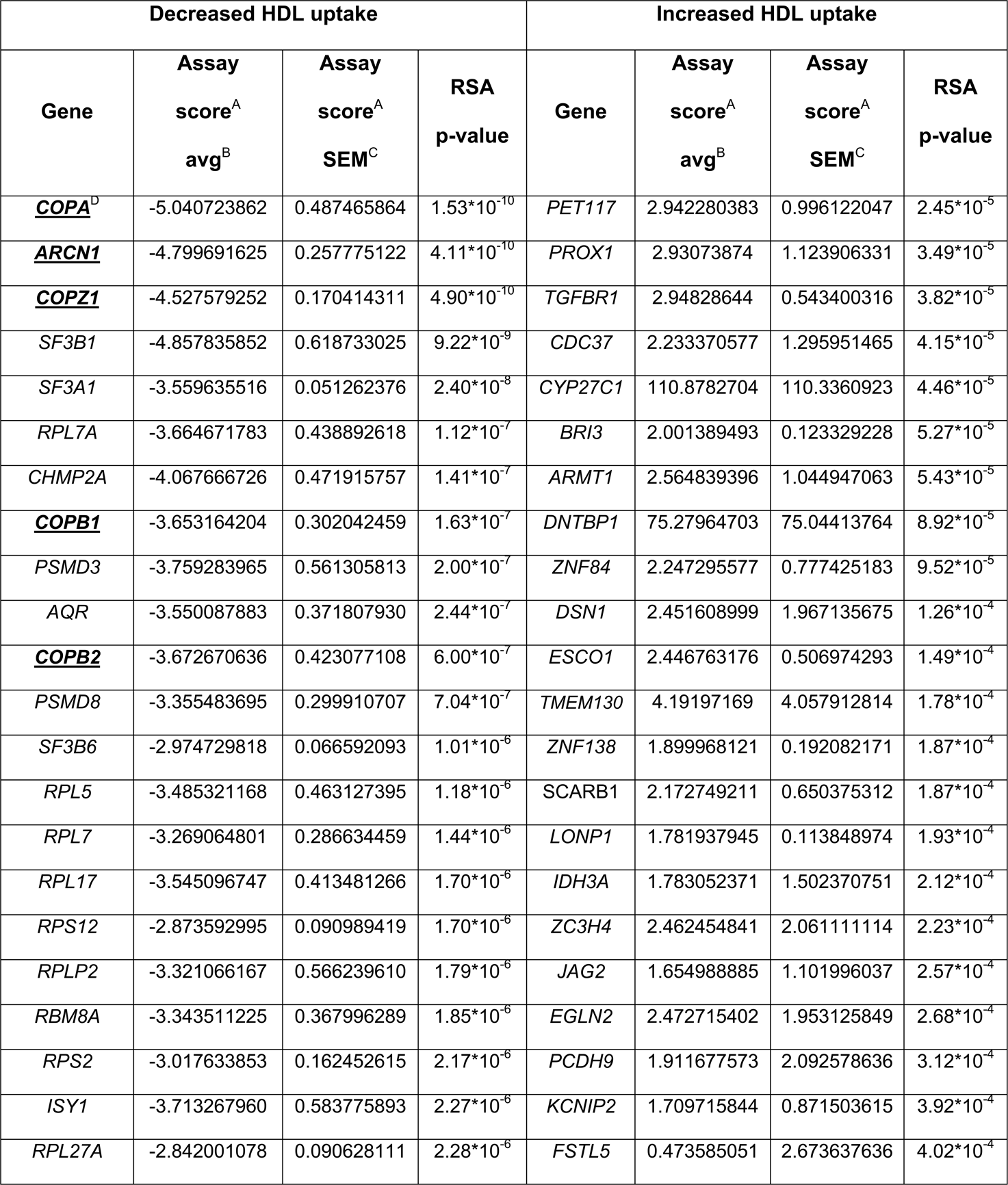
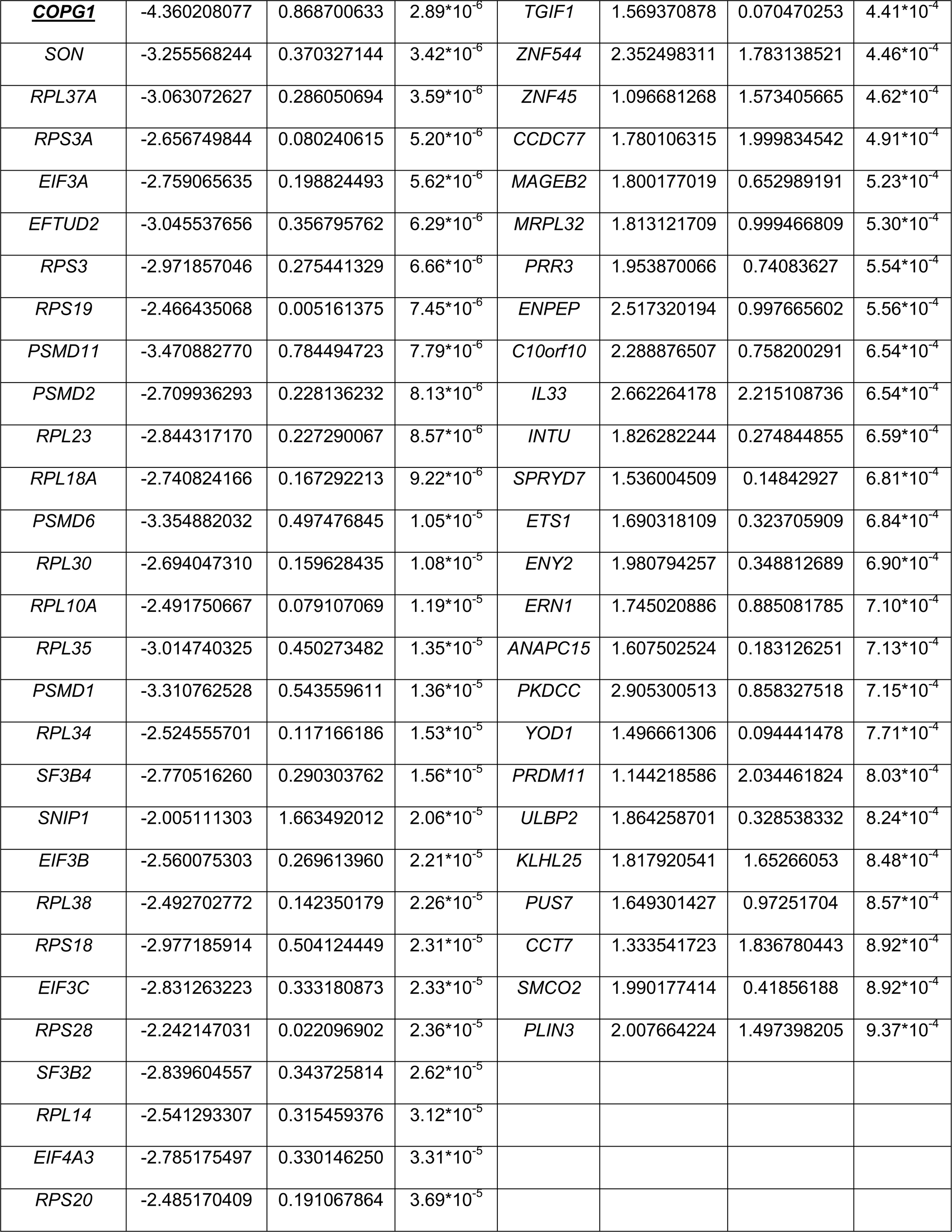
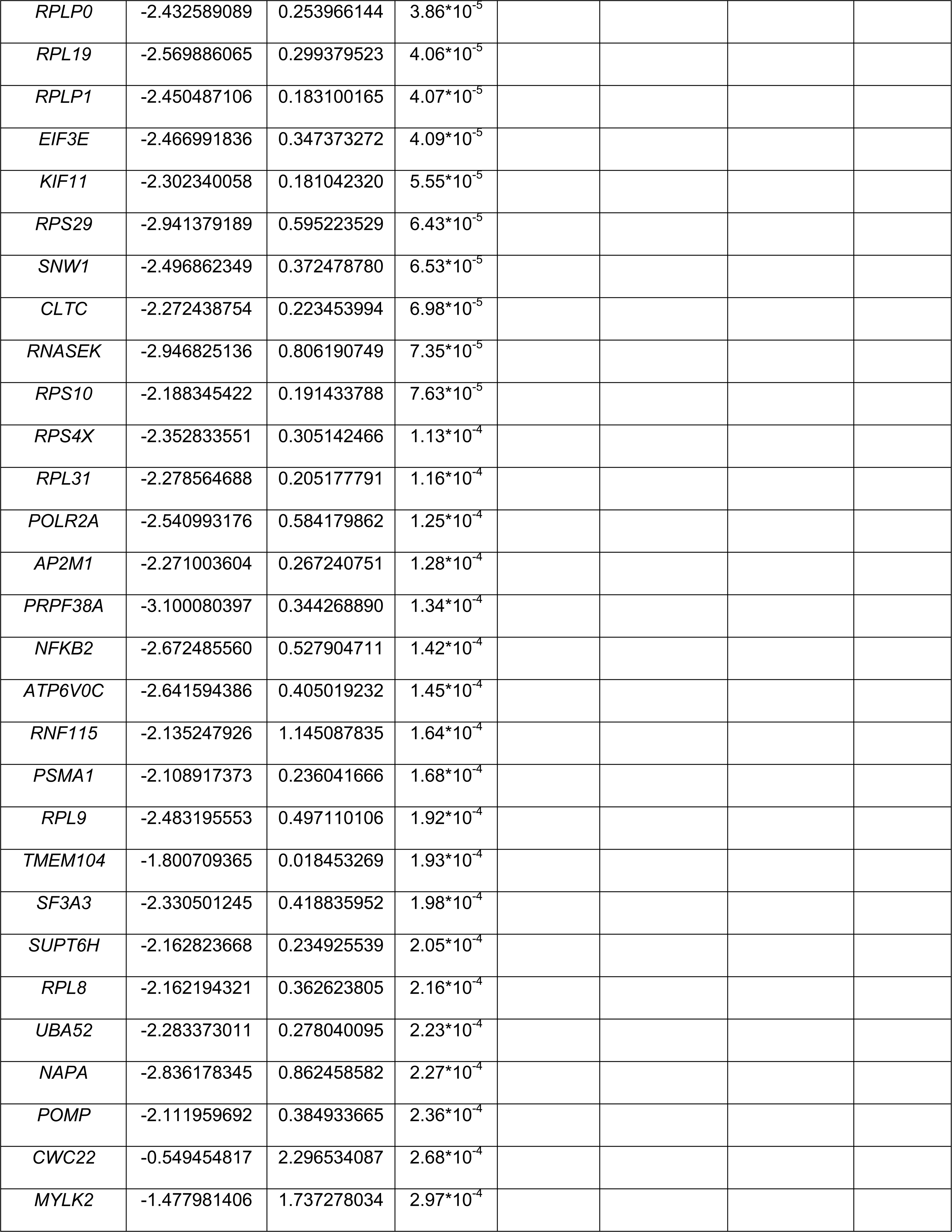
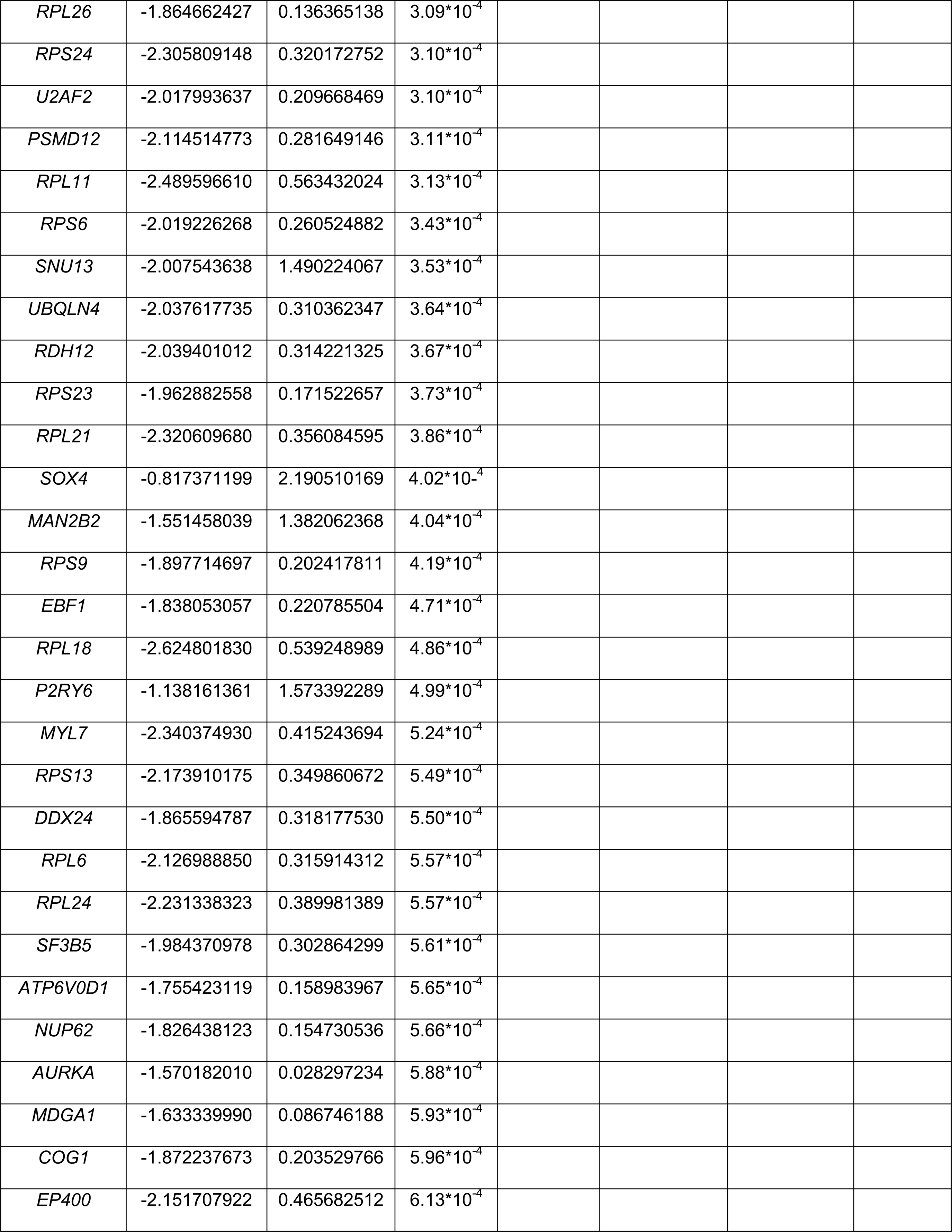
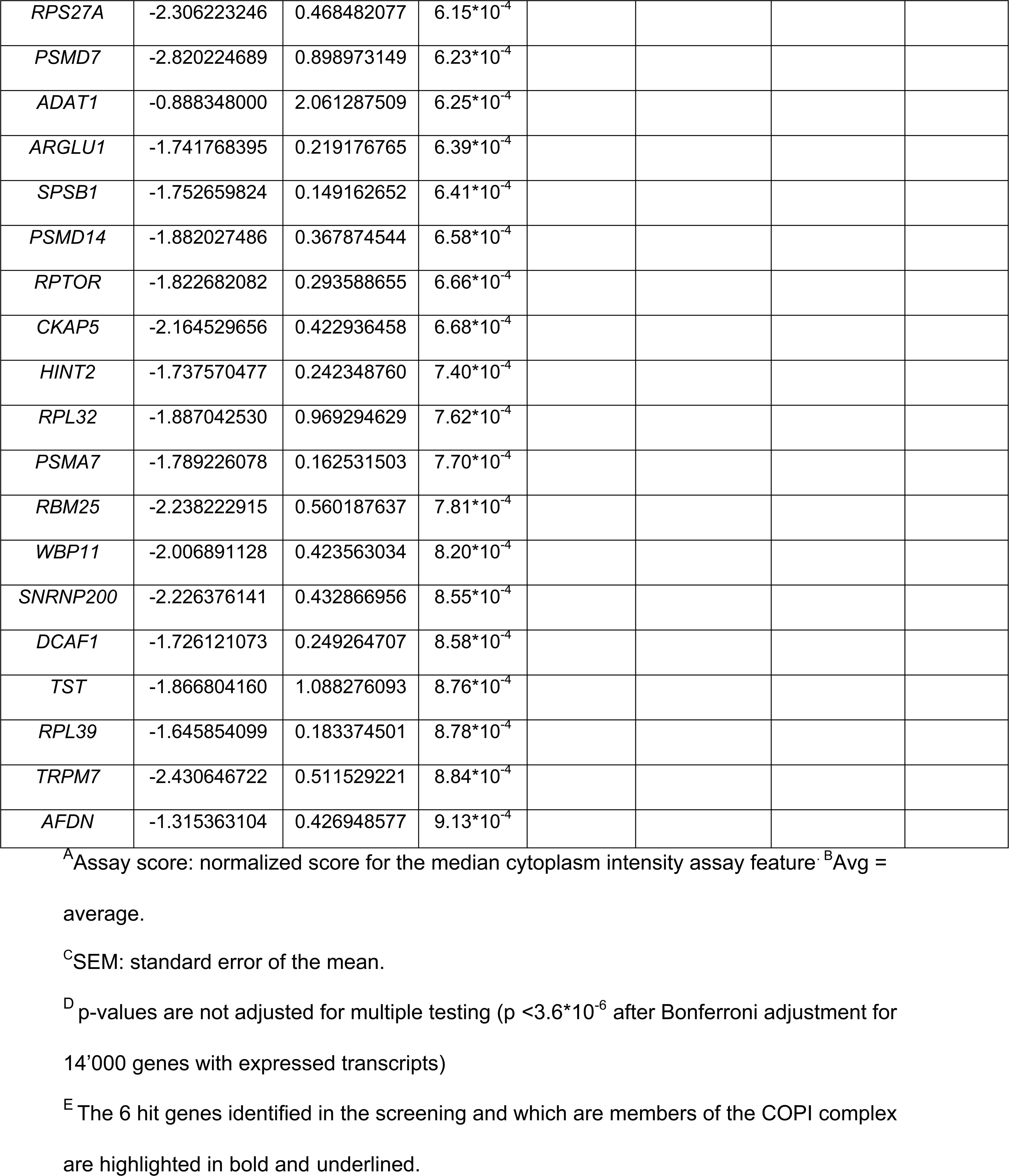
Genes whose loss-of-function in Huh-7 cells cause either a decrease (left column) or an increase (right column) in HDL uptake.

### *In vitro* validation of the COPI complex as a rate-limiting factor in HDL endocytosis

To validate these findings, we utilized pooled siRNAs from a different vendor than the siRNA screening library to knockdown target genes in Huh-7. The knockdown reduced the mRNA expression of the targeted genes by 67% to 90% (supplemental figure S2A) and led to the near disappearance of the encoded proteins (figures 1B-1E). Of note, while non-targeted COPI mRNAs were not suppressed (supplemental figure S2A), the abundance of some non-targeted COPI proteins was substantially decreased, probably because they form complexes with the targeted protein^4^ (figures 1B-1E). For example, the abundance of ε-COP was decreased in the absence of *COPA* or *COPB2,* which form the B subcomplex (figure 1B). Similarly, the abundance of α-COP was decreased by interference with *COPB2* but not *COPE*, in agreement with the dispensable role of ε-COP (figure 1C). The abundance of γ-COP r δ-COP was decreased by knockdowns of genes that encode subunits of the F subcomplex, i.e. *COPB1*, *ARCN1*, *COPG1,* and *COPZ1* (figures 1D and 1E).

We next used flow cytometry to investigate the effects of siRNA interference against each COPI component on the uptake of HDL labelled on the protein moiety (Atto655-HDL) (figures 1F and 1G) or lipid moiety 1,1’-dioctadecyl-3,3,3’,3’-tetramethylindocarbocyanine perchlorate (DiI-HDL) (figures 1H and 1I). Knocking down the indispensable components of the COPI coatomer (*COPA, COPB1, COPB2, ARCN1, COPG1*, or *COPZ1*) reduced Atto655-HDL (figures 1F and 1G), and DiI-HDL (figures 1H and 1I) uptake by at least 75% and 79%, respectively, the latter to a similar extent as knocking down *SCARB1* that was used as positive control for selective lipid uptake from HDL^2^. Conversely, knocking down the dispensable *COPE* as well as the paralogous *COPG2* or *COPZ2* did not alter Atto655-HDL or DiI-HDL uptake. Despite these large effect sizes, when analyzed individually, the differences measured upon interferences with indispensable COPI subunits did not achieve statistical significance after correcting for multiple testing (figures 1F and 1H). However, the decreases became statistically significant when comparing the combined data on the six indispensable COPI subunits versus the combined data on the three dispensable or paralogous COPI proteins (figures 1G and 1I, both p < 0.001). Taken together, the loss of indispensable subunits of the COPI coatomer compromises the uptake of HDL by Huh-7 cells.

### Glycosylation and cell surface expression of SR-BI are compromised upon silencing of different COPI genes

As expected because of its canonical role in selective uptake^2^, the loss of SR-BI decreases the uptake of DiI-HDL into Huh-7 cell (figure 1H). Both in the siRNA screening (table 1) and in targeted replication experiments (figure 2A), we found that the knockdown of *SCARB1* increases the uptake of protein-labeled Atto655-HDL. Interestingly, this increase was not prevented by the additional knockdown of *ARCN1* (figure 2A) underscoring the missing contribution of SR-BI to HDL-holoparticle uptake into Huh7-cells. To explore the reason for the decreased activity of SR-BI in selective lipid uptake, we investigated the effects of lost COPI function on the expression of SR-BI. Silencing of the different COPI genes did not alter SR-BI mRNA or protein levels (supplemental figures 2B and 2C). However the electrophoretic mobility of SR-BI was accelerated upon knockdown of *COPA*, *COPB1*, *COPB2*, *ARCN1*, *COPG1*, and *COPZ1* but unaltered upon knockdowns of *COPE*, *COPG2,* or *COPZ2* (Figure 2B) suggesting an influence on post-translation modification. We hypothesized that the increased mobility of SR-BI reflects decreased glycosylation in response to the loss of COPI proteins. Testing this hypothesis we found that the treatment of Huh-7 cells with neuraminidase (that removes sialic acid residues) alone or in combination with O-glycosidase (that removes O-linked disaccharides) but not with O-glycosidase only, resulted in similar changes of SR-BI’s electrophoretic mobility as observed upon knockdown *of ARCN1*, while treatment with PGnase F (that removes N-linked glycans) led to a much more profound increase of SR-BI’s mobility (figure 2C). In agreement with SR-BI being only N-glycosylated^16^, our results suggest that the knockdown of indispensable COPI genes interferes with the sialylation of SR-BI.

**Figure 2.**
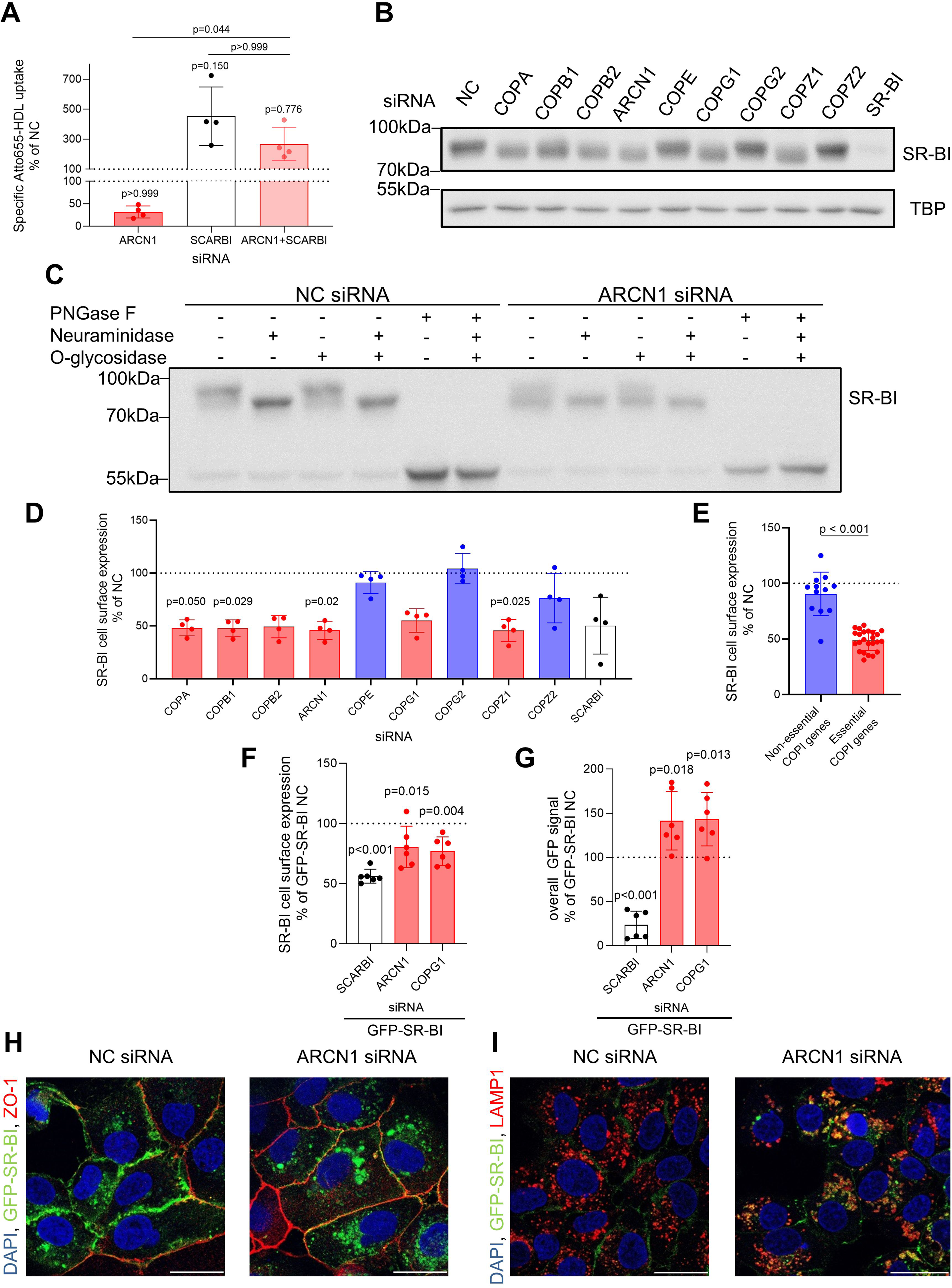
Effect of silencing COPI genes on the HDL-uptake activity, protein expression, glycosylation and localization of SR-BI. Wild type Huh-7 cells (**A**-**E**) or Huh-7 cells overexpressing GFP-SR-BI (**F**-**I**) were transfected with the indicated siRNAs. 72 hours after transfection, the cells were collected and used for functional evaluation. **A**. The uptake of HDL was recorded as described in the legend of figures 1F and 1G. The data were normalized to the non-coding control (NC) and are shown as means ± SD of 4 independent experiments for each condition separately. **B** and **C** show representative Western Blots of SR-BI each out of 3 independent experiments using lysates immediately after harvesting (**B**) or after overnight treatment with the indicated combinations of deglycosylating enzymes (**C**). TATA binding protein (TBP) served as the loading control in (**B**). Figures **D**-**F** show SR-BI cell surface levels of alive Huh-7 cells as determined by flow cytometry with an antibody against an extracellular epitope of SR-BI. The fluorescence emitted by the GFP-SR-BI was recorded as a measure of total cellular SR-BI (**G**). All data were normalized to the non-coding (NC) control and are shown as means ± SD of 4 (**D**,**E**) or 6 (**F**,**G**) independent experiments. **E** compares merged data on the non-dispensable COPI subunits (red bars) and the dispensable or paralogous COPI genes (blue bars). Figures **H** and **I** depict confocal microphotographs on the colocalization of GFP-SR-BI with ZO-1 (**H**) and LAMP1 (**I**). The data shown are representative of 2 independent experiments. The scale bar for both microphotographs is 25 μm. Statistical analysis was performed using Kruskal-Wallis test with Dunn’s multiple comparisons test between the indicated conditions (**A**) or between the NC and each targeting siRNA (**D**), or 1-way ANOVA coupled with Dunnett’s test for multiple comparisons between the NC and each targeting siRNA (**F**,**G**), or two-tailed unpaired t-test between the essential and non-essential conditions (**E**). In (**D**-**G**) only statistically significant differences (p<0.05) are shown.

Both western blotting after cell surface biotinylation and flow cytometry analysis with an anti-SR-BI antibody revealed a strongly reduced cell surface abundance of SR-BI upon knockdown of each indispensable COPI subunit gene similar to the knockdown of *SCARB1* (supplemental figure 3A, figures 2D and 2E). Since western blotting after knockdown of SCARB1 detected no or only traces of SR-BI immunoreactivity both in the total cell lysate and on the cell surface (figure 2B and supplemental figure SA), we consider the residual signal from the flow cytometry analysis as unspecific background (figure 2D). The statistical analysis of the combined data revealed a significant decrease of SR-BI cell surface abundance after knockdown of indispensable COPI genes versus dispensable and paralogous COPI genes (p<0.001, figure 2E).

To corroborate that the loss of COPI genes interferes with the trafficking and cell surface expression of SR-BI, we overexpressed SR-BI with an N-terminal GFP-tag in Huh-7 cells and an empty vector (EV) as the control. Western blot analysis with either an anti-SR-BI or an anti-GFP antibody confirmed the successful overexpression of the construct (supplemental figures S3B and S3C). Cells overexpressing GFP-SR-BI (clone “6” in supplemental figures S3B and S3C) emitted fluorescence and displayed more anti-SR-BI-immunoreactivity on the cell surface than the EV cells (supplemental figures S3D and S3E, respectively). Silencing of *ARCN1* or *COPG1* in Huh-7 cells overexpressing GFP-SR-BI decreased cell surface levels of SR-BI but increased the total GFP signal (figures 2F and 2G, respectively).

We next examined with confocal microscopy the subcellular distribution of the GFP signal in Huh-7 cells overexpressing GFP-SR-BI. Upon silencing of *ARCN1,* the finely granulated GFP signal disappeared from the ZO-1 positive plasma membrane to a great extent, but accumulated in LAMP1-positive late endosomes and lysotracker-positive lysosomes (figure 2H, 2I and supplemental figure S4A-S4C). The GFP signal was co-localized with neither the ER (SEC61A1, supplemental figure S4D) nor the Golgi (GM130, supplemental figure S4E).

### Silencing of COPI genes increases the cell surface abundance of ABCA1 and cholesterol efflux from hepatocytes

In various cell types, cholesterol efflux has been shown to involve endocytosis and re-secretion of HDL^2^. We therefore investigated the effects of lost COPI function on ABCA1 expression and cholesterol efflux. *ABCA1* mRNA levels were significantly decreased by 30% upon silencing of indispensable COPI genes compared to non-essential genes (figures 3A and 3B, p < 0.001). Similarly to SR-BI, knockdowns of essential COPI genes increased the electrophoretic mobility of the ABCA1 band (figure 3C) possibly indicating altered glycosylation of ABCA1. To examine whether the lack of essential COPI coatomer affects the cell surface abundance of ABCA1, we carried out cell surface biotinylation experiments in Huh-7 cells after *ARCN1* knockdown. Silencing of *ARCN1* had no significant effect on total lysate ABCA1 levels but led to a non-significant two-fold increase of ABCA1 abundance on the cell surface (figures 3D and 3E). In agreement with the increased ABCA1 cell surface expression, silencing of COPI genes tended to increase cholesterol efflux from Huh-7 cells to either lipid-free apoA-I (figures 4A and 4B) or HDL (figures 4C and 4D). Co-silencing of *ARCN1* with *ABCA1* (figures 4E and 4F), but neither *ABCG1* nor *SCARB1 (*supplemental figures S5A-S5D) significantly reduced the cholesterol efflux to lipid-free apoA-I as well as HDL, indicating that ARCN1 influences cholesterol efflux by regulating the activity of ABCA1.

**Figure 3.**
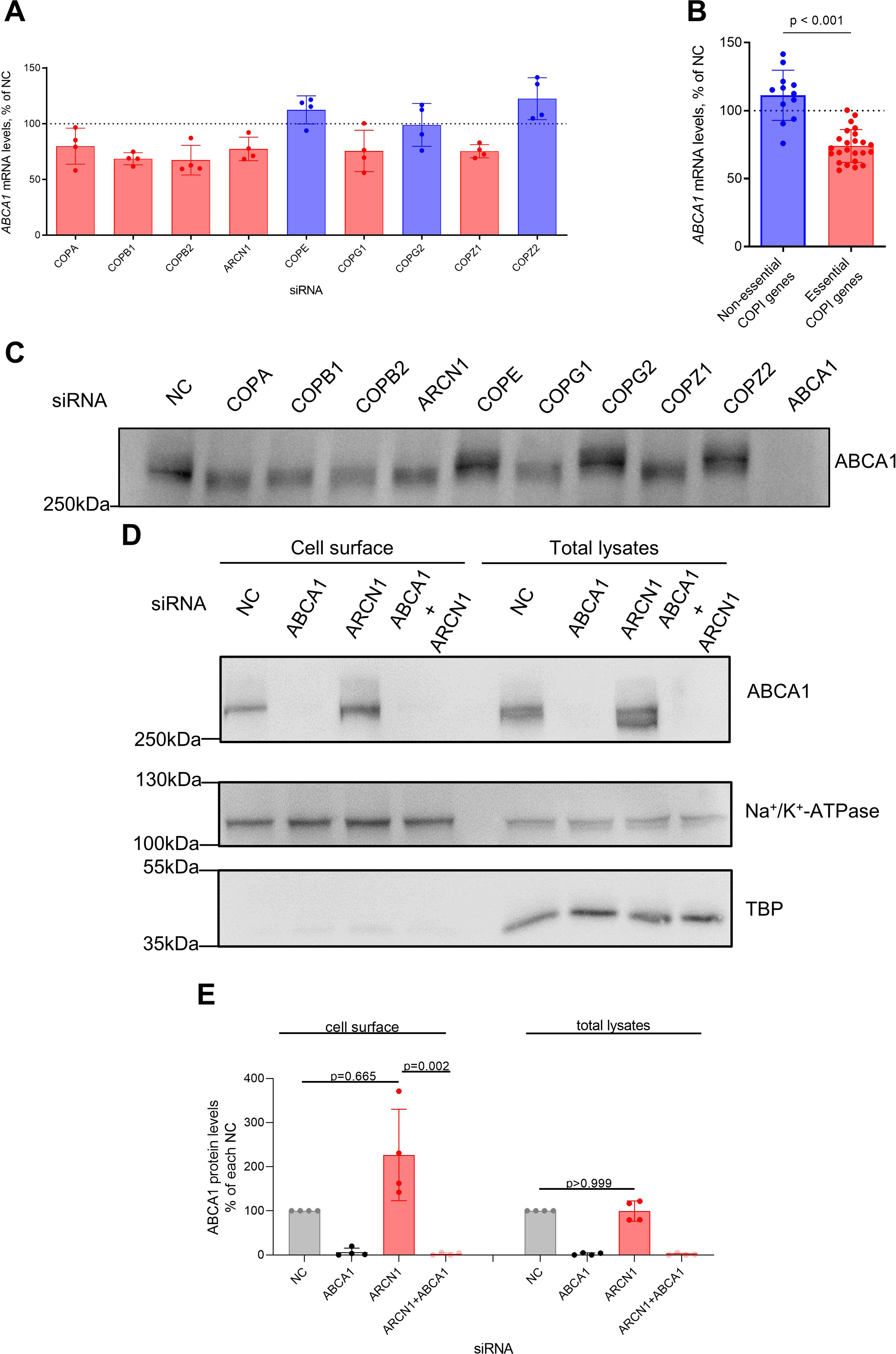
Effect of silencing COPI genes on the expression and the cell surface abundance of ABCA1. Wild type Huh-7 cells were transfected with the indicated siRNAs (**A**-**E**). 72 hours after transfection, the cells were harvested and used for the evaluation of mRNA (**A**,**B**), protein (**C**) and cell surface levels (**D**,**E**) of ABCA1. **A** and **B**. The data were normalized to the non-coding (NC) control and are shown as means±SD of 4 independent experiments. **B** compares the summarized data on the non-dispensable COPI subunits (red bars) with the data on the dispensable or paralogous COPI genes (blue bars). **C** shows a representative Western Blot from 3 independent experiments. For the analysis of ABCA1 expression on the cell surface (**D-E)**, the cells were incubated 48 hours after transfection with DMEM supplemented with 0.5% FBS, 1% P/S and 10 μM T0901317 for 24 hours prior harvesting. The western blots were probed with antibodies against ABCA1, TBP (as a control for intracellular protein expression), and Na^+^/K^+^-ATPase (as a control for cell surface protein expression). For details see methods. **D** shows one representative Western Blot out of 4 independent experiments. **E** shows the respective quantification of the obtained ABCA1 immunoreactive signal normalized either to Na+/K+-ATPase or to TBP. The data are normalized to the non-coding control (NC) and shown as means ± SD of 4 independent experiments. Statistical analysis was performed using either Kruskal-Wallis test with Dunn’s test for multiple comparison between the NC and each targeting siRNA (**A**), or Kruskal-Wallis test coupled with Dunn’s test for multiple comparisons for the indicated conditions (**E**) or two-tailed unpaired t-test (**B**). In (**A**,**B**), only levels of statistical significance with p<0.05 are shown.

**Figure 4.**
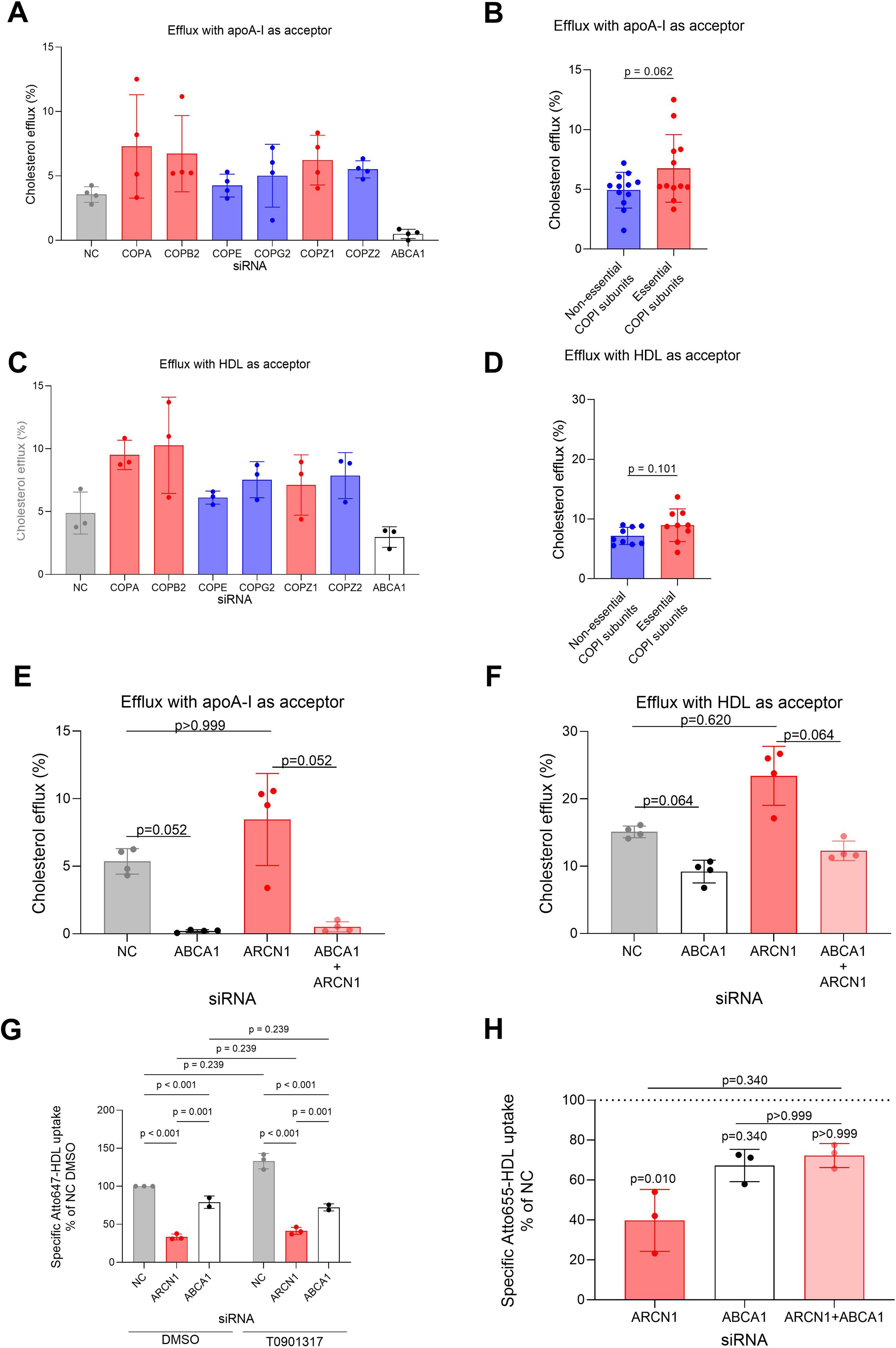
Effects of silencing COPI genes or *ABCA1* on cholesterol efflux and HDL uptake by Huh-7 cells. **A**-**F.** [^3^H]-cholesterol efflux was measured 72 hours after transfection of Huh-7 cells with the indicated siRNAs by using lipid-free apoA-I (**A**,**B,E**) or HDL (**C,D,F**) as the acceptors. The data are shown as means±SD of 4 (**A,B,E,F**) or 3(**C**,**D**) independent experiments. **B** and **D** compare the summarized data on the non-dispensable COPI subunits (red bars) with the data on the dispensable or paralogous COPI genes (blue bars). **G** and **H**. 48 hours post transfection and treatment with 10 μM of the LXR agonist T0901317 for 24 hours (**G**) or 72 hours post transfection (**H**) the cells were incubated with 20 μg/mL Atto647-HDL (**G**) or Atto655-HDL (**H**) for 3 hours. The specific cell association was determined as described in the legend of figure 1. The data are shown as means ± SD of 3 independent experiments. Statistical analysis was performed using either Kruskal-Wallis test coupled with Dunn’s multiple comparisons test between the NC and each targeting siRNA (**A**,**C**) or between the indicated conditions (**E,F,H**) or 2-way ANOVA coupled with Bonferroni’s test for multiple comparisons (**G**) or using two-tailed unpaired t-test (**B**,**D**). In (**A**-**D**) only significant findings with p<0.05 are indicated.

We next investigated whether *ABCA1* contributes to the decreased Atto655-HDL uptake observed upon loss of *ARCN1 (*figures 4G and 4H). Upregulation of ABCA1 by treatment with an LXR agonist increased the uptake of Atto647-HDL by 33%, although not statistically significant (figure 4G). In both unstimulated and stimulated cells, silencing of *ARCN1* and *ABCA1* decreased the uptake of Atto647-HDL by 66.6% and 20.9% and by 68.9% and 45.7%, respectively (p < 0.001 for both, figure 4G). The comparison of residual Atto647-HDL uptake between stimulated and non-stimulated cells after knockdown of either *ARCN1* or *ABCA1* did not reveal any statistically significant difference. When compared to single knockdown of *ABCA*1, the additional knockdown of *ARCN1* did not further decrease the uptake of Atto655-HDL (figure 4H). Taken together our findings suggest a contribution of ABCA1 to the uptake of protein-labeled HDL by Huh-7 cells as well as an interaction of ABCA1 and ARCN1 in this process.

### Loss of COPI genes impairs the secretion of apoA-I

Since hepatic ABCA1 contributes to the biogenesis of HDL, we tested the effects of loss of COPI gene expression on the secretion of apoA-I. Silencing of the essential COPI genes decreased the expression of *APOA1* mRNA by 35% to 57% (figure 5A). The 58% difference between indispensable and non-essential COPI genes was significant (figure 5B, p < 0.001). The secretion of apoA-I was decreased by 41% and 22% upon knockdown of *COPA* (p = 0.045) and *ARCN1*, respectively, but unaltered upon knockdown of *COPG1* (-3%) and increased by 33% upon knockdown of *COPG2* (figure 5C). The comparison of the former three with the paralogous COPG2 revealed significantly decreased apoA-I secretion upon loss of the indispensable COPI genes (figure 5D).

**Figure 5.**
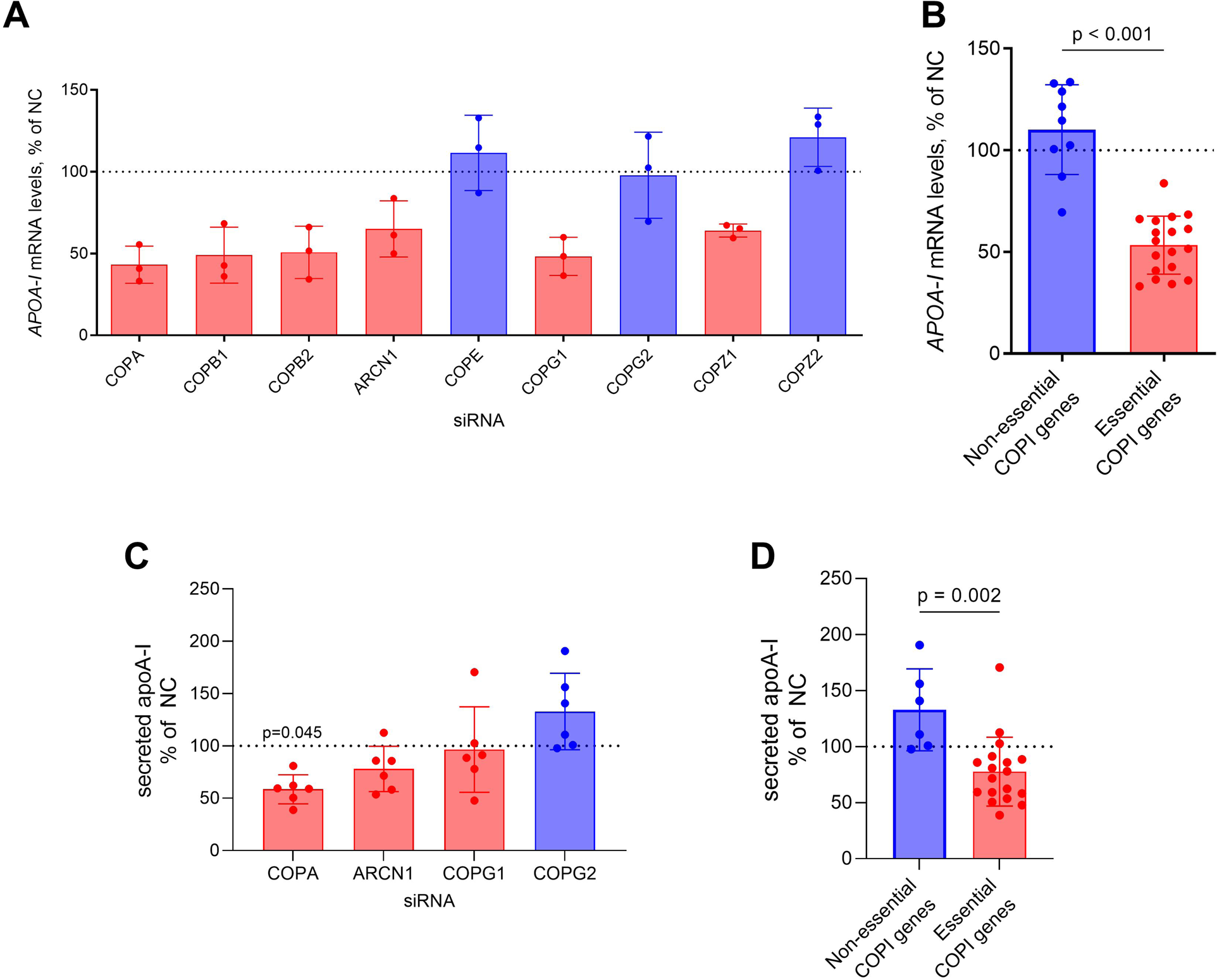
*APOA1* expression (A,B) and apoA-I secretion (C,D) by Huh-7 cells depleted of different COPI genes. Huh-7 cells were transfected with the indicated siRNAs. After 72 hours the cells were collected for quantification of *APOA1* mRNA levels by qRT-PCR (**A**,**B**). For quantification of apoA-I secretion (**C**,**D),** the cells were reseeded 48 hours after transfection. 6 hours later the medium of the cells was replaced by DMEM containing neither phenol red nor FBS. After another 48 hours, both cells and media were collected with the former being counted and the latter being used for the measurement of apoA-I by ELISA. ApoA-I concentrations in the media were normalized to the cell number of each condition. The data are shown as means ± SDs of 3 (**A,B**) or 6 (**C,D**) independent experiments, each normalized to the NC. **B** and **D** compare the data of the non-dispensable COPI subunits (red bars) versus the data of dispensable or paralogous genes (blue bars). Statistical analyses were performed using either Kruskal-Wallis test coupled with Dunn’s multiple comparison test (**A**), 1-way ANOVA coupled with Dunnett’s test for multiple comparisons (**C**) between the NC and each targeting conditions, two-tailed unpaired t-test (**B**) or two-tailed Mann-Whitney test (**D).** Only statistically significant differences (p<0.05) are shown.

### Effect of *ARCN1* on cellular sterol homeostasis

We next investigated the effect of *ARCN1* silencing on the sterol homeostasis in Huh-7 cells in the presence of HDL and under the two conditions that we used for the measurement of HDL uptake (i.e. without LXR agonist) and cholesterol efflux (in the presence of LXR agonist T0901317 and ACAT inhibitor Sandoz 58-035), (supplemental table S2). In agreement with higher HDL uptake and lower cholesterol efflux, cells lacking *ARCN1* had a ∼24% lower content of cholesterol under either condition. However, the content of cholesteryl esters was increased by the knockdown of ARCN1 so that the decrease of cholesterol was caused by the decrease in unesterified cholesterol (supplemental figures S6A and 6B). The increase in cholesteryl esters was not prevented by ACAT inhibition so that the loss of *ARCN1* appears to affect the hydrolysis rather than production of cholesteryl esters (supplemental figure 6A). The lack of *ARCN1* also led to lower cellular contents of most phytosterols (supplemental table S2). As the counter-regulatory response to the decreased HDL uptake and increased cholesterol efflux, the knockdown of *ARCN1* led to higher cellular concentrations of cholesterol precursors, most prominently for dihydrolanosterol in both conditions (+127%, and 85%) and for lanosterol and desmosterol in the condition of stimulated cholesterol efflux (+48%, and +23%, respectively). However, none of the described differences was statistically significant (supplemental table S2).

### Genetic data indicate that the COPI complex regulates HDL-C levels in humans

To explore any limiting effects of the COPI coatomer on HDL metabolism, we tested the associations of 5,821 SNPs in the nine COPI genes with HDL-cholesterol in the most recent aggregated data sets of the Global Lipids Genetics Consortium using >1.65 million individuals^17^. At the Bonferroni adjusted GWAS threshold of p = 9.60*10^-10^, and in line with the reduced HDL uptake into Huh7 cells lacking *ARCN1*, 40 SNPs of *ARCN1* were associated with significantly higher levels of HDL-C (figures 6A) but no difference in triglycerides (supplemental figure S7A). The SNPs are in strong linkage disequilibrium (LD > 0.8). Among them, the intronic variant rs73023350 has the strongest association with HDL-C as well as with gene expression of ARCN1. C*OPB1* harbors four and eleven SNPs, which were associated with significantly higher and lower levels of HDL-C, respectively and one SNP in *COPG2* was associated with lower levels of HDL-C (figure 6A). All five SNPs of *COPZ2* were associated with significantly higher levels of HDL-C as well as lower levels of triglycerides (supplemental figure S7A).

**Figure 6.**
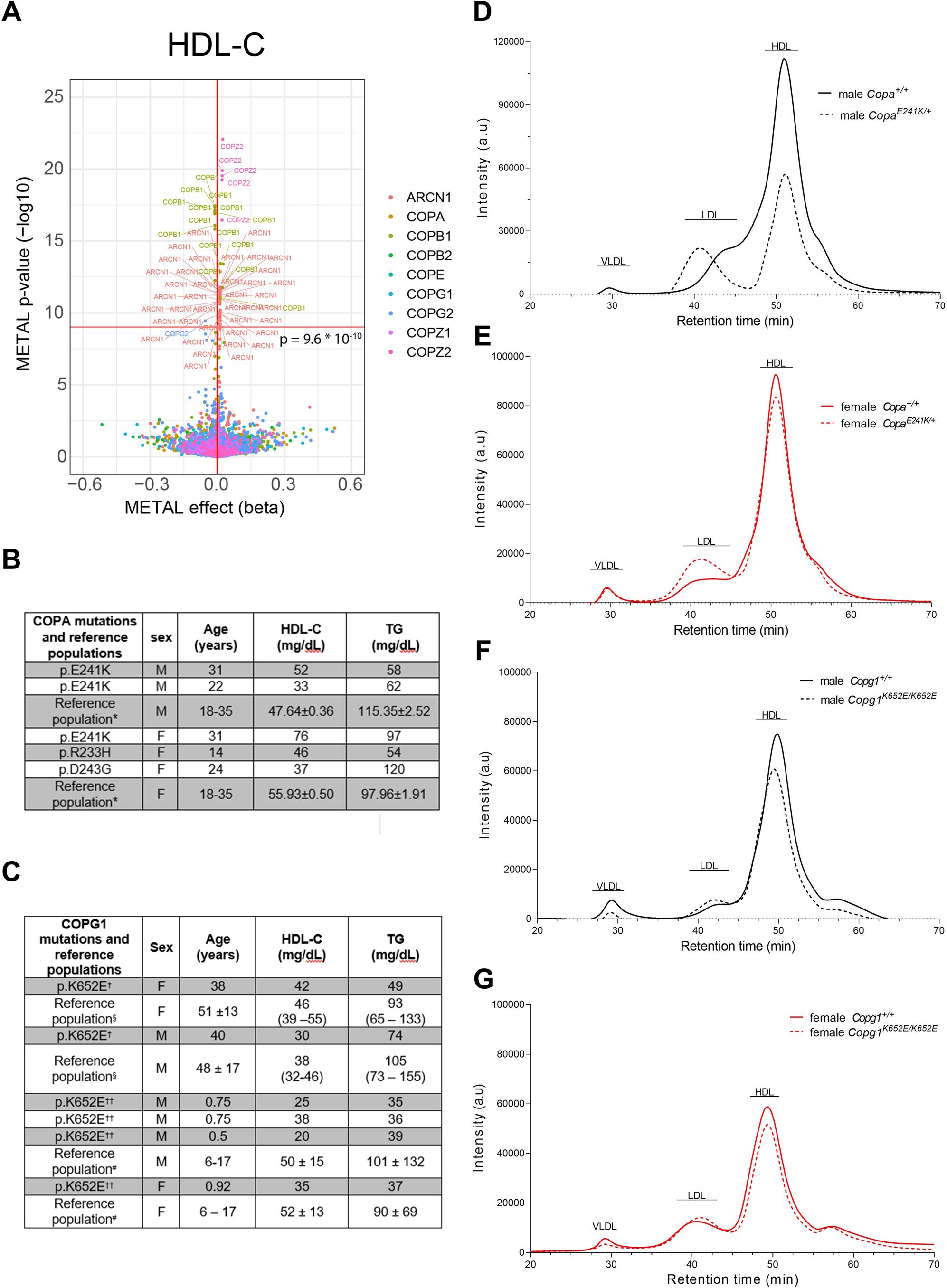
Associations of SNPs (A) or rare pathogenic variants (B-G) of COPI genes with HDL-C levels in humans (A-C) and mice (D-G). **(A)** Volcano plot depicting the association of 5,821 SNPs of the nine COPI genes with HDL-C in 1.65 million individuals aggregated by the Global Lipid Genetics Consortium^17^. The x-axis depicts the effect size (beta) and the y-axis the p-value. The dashed horizontal line marks the threshold of statistical significance after Bonferroni correction for multiple testing on GWAS level (p = 9.60*10^-10^). Tables **B and C** show HDL-C and triglyceride levels in individuals suffering from COPA syndrome^8^ and COPG1 syndrome^9^, respectively. For comparison with the US-American COPA variant carriers (**B**), data of 6127 male and 6157 female participants of NHANES^19^ are presented (*) as means± standard error. For comparison of the heterozygous (†) and homozygous (††) carriers of the rare COPG1 variant (**C**) who originate from Oman, data of adults^21^ (§) and children and adolescents^20^ (#) from the United Arab Emirates are shown for comparison. Data are means ± SD of 485 men and 492 women(§) or 490 boys and 476 girls (#). HDL-C: HDL-Cholesterol; TG: Triglycerides; SD: Standard deviation. Figures **D through G** show the cholesterol distribution among lipoproteins in the pooled plasmas of three male (**D**) and female (**E**) mice heterozygous for *Copa^E241K^* or five male (**F**) and female (**G**) mice homozygous for *Copg1^K652E^* (dashed lines) and the respective control littermates of the same sex (complete lines). Lipoproteins were fractionated by fast protein liquid chromatography column (FPLC). Cholesterol in the fractions was measured by an enzymatic photometric test. In all graphs, the x-axis describes the time at which the different fractions were collected and the y-axis describes the absorbance of the different fractions measured at 505 nm. a.u: arbitrary units.

The analysis of the Whole Exome Sequencing data of more than 394,000 individuals in the UK Biobank^18^ identified 8553 variants in the investigated COPI genes (Bonferroni adjusted threshold p = 5.8*10^-6^). Among them only one rare synonymous variant in *ARCN1* (ENSP00000264028.4: p.Ile510; p = 1.93 * 10^-6^) and one non-synonymous variant in *COPZ2* (ENSP00000480707.1:p.Gly22Arg; p = 4.27 * 10^-6^) were significantly associated with higher HDL-cholesterol (supplemental figure 7B). Of note, only the rare *COPZ2* variant was also significantly associated with lower triglycerides (p = 5.8 * 10^-6^) supplemental figure 7C).

Finally, we investigated the plasma lipids of five American patients carrying heterozygous pathogenic variants in *COPA* causing a type I interferonopathy^8^ (figure 6B) and four children homozygous for the pathogenic variant p.K652E in *COPG1* causing combined immunodeficiency as well as their heterozygous parents^9^ (figure 6C). Compared to adults from the NHANES reference population^19^, the HDL-C levels of the COPA syndrome patients were either normal or rather low except for one female carrier of the COPA(p.E241K) variant who presented with high levels of HDL-C (figure 6C). Because of their origin from Oman, we compared the HDL-C levels of the homozygous *COPG1* variant carriers and their heterozygous parents with those of populations in the United Arab Emirates^20,21^. HDL-C levels of affected children were rather low while those of their parents were within the normal ranges for the respective control populations (figure 6C).

FPLC profiling of lipoproteins in plasmas of mice with heterozygosity for the *Copa^E241K^* (*Copa^E241K/+^*) mutation (figures 6D and 6E) or homozygosity for the *Copg1^K652E^* mutation (*Copg1^K652E/K652E^*) (figures 6F and 6G), which mimic the immunological phenotypes of the COPA and COPG1 syndromes described above^9,22^, also revealed lower levels of HDL-C in both *Copa^E241K/+^*and *Copg1 ^K652E/K652E^* mice of both sexes compared to wild type mice (figures 6D to 6G). In summary, HDL-C levels were normal or rather low in both the patients and the mouse models of the COPA and COPG1 syndrome.

## Discussion

The molecular mechanism of HDL holoparticle removal by the liver is an unresolved part of HDL metabolism. Its elucidation is not only of biological interest but may also have clinical relevance. Since LDL-R dysfunction impairs LDL removal and thereby leads to LDL-hypercholesterolemia, disturbed HDL removal is expected to increase HDL-C levels. However, it is uncertain how this will affect the risk of ASCVD. HDL-C levels are inversely associated with the risk of ASCVD, however only within the range of the lower 6 or 7 deciles^1^. Moreover, large epidemiological studies have found high HDL–C levels associated with increased risks of total and cardiovascular mortality^1^. Also indicating a detrimental impact of high HDL-C levels due to impaired HDL removal, the plasma concentration of inhibitory factor IF1, which inhibits ecto-F1ATPase^2,3^ and thereby HDL holoparticle uptake, is associated inversely with both HDL-C and a history of myocardial infarction^23^. Likewise, defective selective lipid uptake due to variants of *SCARB1* are associated with increases of both HDL-C and risk of ASCVD^24^. Molecular understanding of HDL removal may help to resolve the question of whether the blockage of HDL removal is beneficial by maintaining potentially protective particles in the circulation or deleterious, for example by delaying reverse cholesterol transport or increasing the lifetime of HDL and thereby the likelihood of potentially harmful particle modifications.^1^

To identify genes that limit the uptake of HDL into hepatocytes, we performed a genome-wide RNA interference screen. This approach was already successfully undertaken by our and other labs to identify regulators of hepatic or endothelial LDL uptake^11,25,26^ but, to the best of our knowledge, has never been applied to HDL. None of the 128 genes identified by our screen to limit the uptake of HDL by Huh-7 cells encoded for a protein that was previously suggested to limit HDL uptake (vigilin^27^, HDL binding protein 2^28^, CD36^29^, ecto-F1ATPase^3^, P2Y13^3^, megalin^30^, cubilin^30^ or any *bona fide* endocytic receptor). The knockdown of *P2YR6*, which encodes the purinergic receptor P2Y6 is noteworthy, because the activation of purinergic receptors P2Y13 and P2Y1 by binding of apoA-I to ectoATPase and subsequent ADP generation was previously shown to promote the HDL holoparticle uptake into hepatocytes and endothelial cells, respectively, by an as yet unresolved endocytic pathway^3^. However, the metabolic effects of P2Y6 have been characterized in skeletal muscle, adipocytes, and hypothalamus rather than in liver^31^, possibly because, although expressed, its activation in Huh-7 and HepG2 cells did not result in any Ca^2+^ mobilization^32^, a typical second messenger response to the activation of purinergic receptors. Also of note, the knockdown of *SCARB1*, which encodes the mediator of selective lipid uptake from HDL^2,24^, increased the uptake of HDL holoparticles.

Our screening identified six of the seven canonical components of the COPI coatomer as limiting factors of HDL uptake, namely *COPA, COPB1, COPB2, ARCN1, COPG1,* and *COPZ1.* By using other siRNAs and flow cytometry as an alternative recording method we confirmed that the loss of each of these six indispensable components of the COPI coatomer results in reduced HDL holoparticle uptake into Huh-7 cells. The only component not identified as contributing by our screening *– COPE –* was also not limiting the uptake of HDL in our targeted verification experiments. Of note, *COPE* is the least evolutionary conserved COPI subunit, and its main role is to stabilize COPA^13^. In the mutant CHO cell line ldlF, the lack of *COPE* interferes with the trafficking of cargo at the non-permissive temperature (39.5°C)^33^. All our HDL uptake experiments were carried out at lower temperature (37°C) although still higher than the permissive one (34°C), suggesting that under these conditions COPE function might be dispensable for the function of the COPI complex. The two other COPI genes *COPG2* and *COPZ2* which did not limit HDL uptake either in the screening or in the validation experiments are paralogs of *COPG1* and *COPZ1,* respectively. Their functions are not well understood but do not appear to limit the functionality of the COPI coatomer^14,15^.

We identified SR-BI, ABCA1, and apoA-I as three targets, whose gene expression (*APOA1* and *ABCA1* but not *SCARB1*), cellular localization (SR-BI and ABCA1), functionality (SR-BI and ABCA1), or secretion (apoA-I) are altered by the loss of COPI subunits and hence coatomer function, but with directionally different effects. Compared to the knockdown of non-essential COPI genes, the interference with indispensable COPI genes led to a significant 30 to 50% reduction in the mRNA expression of *APOA1* and *ABCA1* but no significant change in *SCARB1* expression. The lower mRNA expression of *APOA1* may contribute to the lower apoA-I secretion observed here. However, Western Blotting analysis did not indicate any obvious differences in the total protein levels of SR-BI or ABCA1. Mechanistically, it is known that the COPI coatomer contributes to the retrograde transport of transcription factors or their activators from the Golgi to the ER, for example of uncleaved SREBP-SCAP complexes or the ARA160 coactivator of the androgen receptor^34,35^. It may therefore be that the various transcription factors and cofactors contributing to gene expression of *APOA1, ABCA1 and SCARB1* genes differ by their susceptibility to disturbances of retrograde transport.

In addition to retrograde transport from the Golgi to the endoplasmic reticulum, the COPI coatomer plays an important role for the transport of proteins through the Golgi cisternae, where glycosylation of cargo proteins takes place^4^. Of note, a *de novo* mutation in *ARCN1* was previously identified as the cause of a transient N-glycosylation deficiency during episodes of acute illness^36^. In our hands, SR-BI and ABCA1 showed altered electrophoretic mobility upon knockdown of all six indispensable COPI genes but not the three dispensable genes. By *in vitro* treatment with neuraminidase, we mimicked the altered mobility of SR-BI. Moreover, the altered electrophoretic properties of SR-BI upon loss of COPI genes resembled those previously found upon treatment with sialidase or substitution of the N-glycosylated amino acid residues of SR-BI^16^. As SR-BI is known to be only N-glycosylated^16,37^, we conclude that the loss of each indispensable COPI subunit interferes with the sialylation of SR-BI’s N-glycans. Likewise, the slightly increased electrophoretic mobility of the band corresponding to ABCA1 is also in agreement with defective N-glycosylation. In this regard, it is noteworthy that the knockdown of *ARCN1* enhanced the cholesterol efflux by ABCA1 which is N-glycosylated^38^, but not ABCG1, which lacks any glycosylation^39^. ApoA-I proteoforms however include only a quantitatively minor fraction of a digycan-proteoform, with one mole of Hex and HexNAc^40^ each, which cannot be distinguished from the unglycosylated proteoforms by SDS-PAGE.

The defective N-glycosylation appears to have opposite effects on the trafficking of SR-BI and ABCA1 to the cell surface. While SR-BI levels are reduced at the cell surface, possibly by lysosomal degradation as indicated by our microscopic co-localization experiments, the cell surface abundance of ABCA1 is increased. The localization in the lysosomes rather than in the Golgi indicates that the compromised COPI function and sialyation of SR-BI interferes with the cycling of SR-BI between its two prevailing cellular pools in the plasma membrane and endosomes/lysosomes^41^, for example by the Wiskott Aldrich Syndrome protein and scar homologue (WASH) complex^42^. The loss of function of SR-BI and the resulting decrease in selective lipid uptake explains the decreased uptake of the fluorescent lipid DiI^43^, but not the decreased uptake of the fluorescently labeled protein moiety analyzed by both our screening and validation experiments. Rather by contrast, knockdown of *SCARB1* was found by us to promote the uptake of atto655-labeled HDL, ie. HDL holoparticles, both in our screening and in targeted experiments. The missing effect of lost SR-BI on HDL-holoparticle uptake by Huh7 cells is in line with data from *in vivo* and *in vitro* data from mice and primary hepatocytes with *Scarb1* knock-out, but in disagreement with findings in endothelial cells and non-hepatic cancer cells ^2,29^. As discussed by us in detail previously^2^, the cell specific role of SR-BI for HDL holoparticle uptake may depend on differential splicing or interaction with co-receptors or downstream signaling molecules.

In parallel to enhancing the cell surface abundance of ABCA1, the knockdowns of the essential COPI genes led to a modest increase in cholesterol efflux from Huh-7 cells, which did not reach statistical significance. However, the more pronounced increase of cholesterol efflux in the presence of lipid-free apoA-I compared to HDL and the prevention of the increase by interference with *ABCA1* but not *ABCG1* or *SCARB1* indicates some functional consequence of the higher cell surface abundance of ABCA1. Of note, these findings contrast with previous reports of reduced cholesterol efflux from THP1 macrophages upon knockdown of *COPB1*^44^ or inhibition of ARF1 with brefeldin A or depletion of brefeldin A-Inhibited Guanine Nucleotide-Exchange Protein (BIG1)^45,46^. Differences in cell types and experimental protocols may explain the discrepant findings. Classically, ARF1 is considered as a limiting factor for the formation of COPI coatomers^47^, but it is increasingly recognized that ARF1 also plays important roles in the regulation of exocytosis, endo-lysosomal trafficking and other coatomer-independent actions^48^. Thus, brefeldin A and BIG1 depletion may compromise ABCA1 trafficking and cholesterol efflux also independently of COPI function. The gain of ABCA1 function may contribute to the decreased HDL uptake upon loss of COPI function because ABCA1-deficient macrophages of Tangier disease patients show defective retroendocytosis with enhanced uptake and degradation of HDL^2^. However, in contrast to this hypothesis but in agreement with findings in RAW264.7 macrophages^49,50^, the knockdown of *ABCA1* led to a decrease in HDL uptake if compared to the non-coding control. LXR-mediated induction of ABCA1 expression slightly increased HDL uptake but not in cells depleted of either ABCA1 or ARCN1. This suggests some involvement of ABCA1 in the reduced HDL uptake upon loss of COPI function.

The decreases in HDL particle uptake and selective lipid uptake as well as the increased cholesterol efflux upon loss of COPI function, would deprive Huh-7 cells from cholesterol if not compensated by an increase in *de novo* synthesis. In fact, upon knockdown of *ARCN1* and incubation with HDL with or without LXR activation, the cellular concentrations of total and unesterified cholesterol in Huh-7 cells were decreased while the cellular concentrations of biosynthetic cholesterol precursors and cholesteryl esters were increased. Cholesteryl esters remained increased upon loss of ARCN1 even in the presence of an ACAT inhibitor, suggesting disturbed cholesteryl ester hydrolysis rather than enhanced cholesterol ester formation in the absence of functional COPI coatomers. In view of the important role of cholesterol for the regulation of morphogenesis, it is interesting to note that *de novo* mutations in *ARCN1* have been identified as the cause of craniofacial dysplasia and microcephaly^51^, i.e. similar phenotypes as observed in Smith-Lemli-Opitz syndrome^52^, which is caused by defects in cholesterol biosynthesis.

By limiting HDL holoparticle uptake as well as SR-BI mediated selective lipid uptake and by promoting cholesterol efflux, the loss of COPI coatomer function is expected to increase HDL-C levels, while the decreased apoA-I secretion could counteract these effects. Our analysis of the GWAS data of more than 1.65 million persons published by GLGC^17^ identified several SNPs in *ARCN1* and *COPB1* to be associated with higher plasma levels of HDL-C. Earlier studies in smaller populations had reported significant associations of *COPB1* and *COPG1* with HDL-C. However, SNPs in *COPZ2* are also associated with HDL-C, although loss of *COPZ2* affects neither the glycosylation of SR-BI and ABCA1, nor the HDL particle uptake, nor the cholesterol efflux. It thus appears that *COPZ2* regulates HDL metabolism by other mechanisms. In view of the metabolic interaction of HDL and triglyceride-rich lipoproteins it is interesting to note that in contrast to the SNPs of *ARCN1* or *COPB1*, all SNPs as well as a rare variant of *COPZ2* are associated with both higher levels of HDL-C and lower levels of triglycerides. It may hence be, that *COPZ2* regulates HDL metabolism indirectly via the metabolism of triglyceride-rich lipoproteins.

In contrast to the results in these population-based genetic studies, human carriers of rare variants in COPA and COPG1 which cause inherited immunopathological syndromes, as well as the homologoous genetic mouse models, both have normal or rather low plasma levels of HDL-C. This discrepancy may be due to specific structure-function relationships of the immunopathogenic COPA and COPG1 variants, for example by having stronger inhibiting effects on apoA-I secretion and hence the production of HDL than on the catabolism of HDL by SR-BI and/or HDL holoparticle uptake. Moreover, disturbed COPI function may also affect other regulators of HDL metabolism which were not investigated here. In this regard, it is important to note that the rather low plasma levels of triglycerides in both humans and mice carrying variants of *COPA* or *COPG1* argue against any secondary lowering of HDL-C in response to hypertriglyceridemia, which is considered as the basis of low HDL-C in many individuals of the general population^1^.

In conclusion, we identified components of the COPI complex as limiting factors of important steps in HDL metabolism in hepatocytes and, probably thereby, determinants of HDL-C levels in the human population.

## Data Availability

All data produced in the present study are available upon reasonable request to the authors

## Nonstandard Abbreviations and Acronyms

RNAi: RNA interference
GWAS: Genome wide association study
WES: Whole exome sequencing
FPLC: Fast protein liquid chromatography
TLC: Thin layer chromatography
GC-MS: Gas chromatography mass spectrometry
LDL-C: Low-density lipoprotein cholesterol
HDL-C: High-density lipoprotein cholesterol
TG: Triglycerides
FC: Free cholesterol
CE: Cholesterol ester
*APOA1*, apoA-I: Apolipoprotein A1
COPI: Coat protein I
*COPA*, α-COP: COPI coat complex subunit alpha
*COPB1*, β-COP: COPI coat complex subunit beta 1
*COPB2*, β’-COP: COPI coat complex subunit beta 2
*ARCN1*, δ-COP: COPI coat complex subunit delta
*COPE*, ε-COP: COPI coat complex subunit epsilon
*COPG1*, γ-COP: COPI coat complex subunit gamma 1
*COPG2*, γ’-COP: COPI coat complex subunit gamma 2
*COPZ1*, ζ-COP: COPI coat complex subunit zeta 1
*COPZ2*, ζ’-COP: COPI coat complex subunit zeta 2
*SCARB1*, SR-BI: Scavenger receptor class B type I
ABCA1: ATP binding cassette subfamily A member 1

## Acknowledgements and Sources of Funding

A.v.E.’s team was funded by the Swiss National Science Foundation (31003A-160126, 310030-185109), the 7^th^ Framework Program (FP7) granted by the European Commission (“TransCard” 603091), and the Swiss Systems X program (2014/267 (MRD) HDL-X).

G.P. received funding from the University of Zurich (Forschungskredit, FK-20-037).

P.Z. received funding awards from the Swiss Atherosclerosis Society (AGLA) and the D•A•CH-Gesellschaft Prävention von Herz-Kreislauf-Erkrankungen. A.T.H.’s team was supported by European Commission (“TransCard”, 603091). S.E.H. received grants RG3008 and PG008/08 from the British Heart Foundation, and the support of the UCLH NIHR BRC.. Flow cytometry was performed with equipment of the flow cytometry facility, University of Zurich.

## Disclosures

The RNAi screening was performed at the ETH ScopeM facility (R.M., M.S., S.F.N., A.J.R, S.S.). As by contract, one third of the service costs was paid by the TransCard project grants of A.v.E to cover part of the costs for personnel, infrastructure, and maintenance. S.E.H. has worked as a consultant for Verve and is the Chief Scientific Officer of a UCL spin-out company StoreGene that offers to clinicians genetic testing for patients with FH.

## Supplemental Materials

Materials & Methods

Online Figures S1 to S7

Online Tables S1 to S4

Supplemental references 52 to 59

## Notes

### Competing Interest Statement

The authors have declared no competing interest.

### Funding Statement

Swiss National Science Foundation (31003A-160126 and 310030-185109)
7th Framework Program (FP7) granted by the European Commission (TransCard 603091)
Swiss Systems X program (2014/267 (MRD) HDL-X)
University of Zurich (Forschungskredit FK-20-037)
Swiss Atherosclerosis Society (AGLA)
DACH-Gesellschaft Praevention von Herz-Kreislauf-Erkrankungen
grants RG3008 and PG008/08 from the British Heart Foundation

### Author Declarations

Institutional Review Boards (IRB) for the protection of human subjects of the University of California in San Francisco (IRB protocol 10-02467) and Boston Childrens Hospital (IRB protocol 04-09-113R).

