## supplemental methods, figures and tables for "The COPI coatomer regulates several steps of HDL metabolism"

#### Table of Contents


| | |
|---|---|
| **Supplemental Methods** | **Pages 2-10** |
| **Supplemental figures** | **Pages 11 -17** |
| Supplemental figure S1 | Page 11 |
| Supplemental figure S2 | Page 12 |
| Supplemental figure S3 | Page 13 |
| Supplemental figure S4 | Page 14 |
| Supplemental figure S5 | Page 15 |
| Supplemental figure S6 | Page 16 |
| Supplemental figure S7 | Page 17 |
| **Supplemental tables** | **Page 18-21** |
| Supplemental table S1 | Pages 18 plus separate csv file with complete data |
| Supplemental table S2 | Page 19 |
| Supplemental table S3 | Page 20 |
| Supplemental table S4 | Page 21 |

### Supplemental Materials and Methods

#### Cell culture

Huh-7 cells (cat. JCRB0403, JCRB Cell Bank, Ibaraki, Japan) were cultured in normal growth medium, comprised of Dulbecco's Modified Eagle's Medium (DMEM) (cat. D-5796, Sigma-Aldrich, St. Louis, USA) supplemented with 10% fetal bovine serum (FBS), (cat. 10500056, Thermo Fisher Scientific, Waltham, Switzerland) and 100 U/mL each penicillin/streptomycin (P/S) (cat. 15140122, Thermo Fisher Scientific).

#### Isolation and labeling of HDL

Human plasmas of normolipidemic donors were purchased from Blutspendedienst Zurich. HDL (1.063<d<1.21 kg/L) was isolated by sequential ultracentrifugation at approximately 257,000 g, 15°C for ~16 hours using the OptimaTM L-90K Ultracentrifuge and the Type 70 Ti fixed angle rotor (Beckman Coulter, Brea, USA)[11]. Density was adjusted with potassium bromide by using a KRUESS DS7000 densitometer (Kruess). To avoid any oxidation, EDTA was added to a final concentration of 30 mM.

HDL was labeled with Atto655 (cat. AD655-35, Atto-Tec, Siegen, Germany) or Atto647 (cat. AD647-35, Atto-Tec) as reported previously[53]. For DiI-HDL uptake experiments, HDL was labeled with 1,1'-dioctadecyl- 3,3,3',3'-tetramethylindocarbocyanine perchlorate (DiI) (cat. D282, Invitrogen). Briefly, on day 1, DiI was dissolved in DMSO to a final concentration of 3 mg/mL. Thereafter, 10 mg HDL was mixed with 10.5 mL of human lipoprotein deficient serum (LPDS) and 263 µL of DiI. After overnight incubation at 37°C in the dark, DiI-HDL was isolated by ultracentrifugation after adjusting density to1.21 g/L with KBr. Isolated DiI-HDL was extensively dialyzed (0.3 mM EDTA and 0.15 M NaCl, pH= 7.4) at 4°C protected from light.

#### siRNA genome-wide screening

The genome-wide siRNA screening for genes limiting HDL uptake into Huh-7 cells was performed together with our previous screening for genes limiting LDL uptake[11]. Briefly, each of the 21,584 human genes were targeted by three unique non-overlapping siRNAs contained by the Ambion Silencer Select Human Genome siRNA library V4 (cat. 4397926, Thermo Fischer Scientific). As the internal controls, each of the following Ambion Silencer Select siRNA oligonucleotides (Thermo Fischer Scientific) were plated in four replicate wells in each of the 192 assay plates: anti-PLK1 (assay ID s448, cat. 4390824) and the Silencer™ Select Negative Control No. 1 siRNA (cat. No. 4390843). Since no endocytic HDL receptor is known[2] and our explorative pilot experiments excluded candidate genes (*e.g. SCARB1, PDZK1*) , we could not include any condition to control for specificity such as *LDLR* in our previous screen for genes regulating LDL uptake[11]. To determine the overall signal-to-noise ratio of our assay, cells transfected with a non-targeting siRNA were incubated in the absence of fluorescent lipoproteins (background control). 72 hours after transfection cells were exposed to a mixture of Atto655-HDL and Atto594-LDL[11] (see also section "Isolation and labeling of HDL" described above) in DMEM, at a final concentration of 33 µg/mL each. After incubation for 4 hours at 37°C, 5% CO2, the cells of each well were washed with PBS, fixed in an isotonic 2% paraformaldehyde solution containing 20 µg/mL of Hoechst 33258 (cat. 861405, Sigma Aldrich) washed again with PBS and stored in a 0.05% solution of sodium azide in water. For each assay plate, two imaging datasets were collected by two twin ImageXpress micro HCS microscopes (Molecular Devices, San José, USA). For cell counting, the whole surface of each well was captured at 4x magnification in the DAPI channel. Additionally, 9 tiled sites in each well were then acquired with a 20x objective in the DAPI and Cy5 channel to allow for the final image analysis. After acquisition, image segmentation and the subsequent image analysis were performed using Cell Profiler (http://cellprofiler.org/, The Broad Institute, Cambridge, USA). The annotation of the Ambion library used in the screening was done using the NCBI.geneID history published on 14$^{th}$ March 2016. Optimal transfection efficiency was confirmed across all screening plates by determining the amount of cell toxicity upon knockdown of the essential gene *PLK1*[11,54]. As for our previous LDL screen[11], we applied the Redundant siRNA Activity (RSA) analysis[11] to data from the best performing assay feature, namely median cytoplasm intensity for the identification of hit genes. Z'-factor values[11] for median cytoplasm intensity in each assay plate for the background without fluorescent HDL (median 0.79, interquartile range [IQR] 0.68 to 0.90 ) indicated excellent signal-to-noise ratio.

**siRNA transfection in validation experiments**

Huh-7 cells were reverse transfected with either siRNAs against the indicated target genes (*COPA*, *COPB1*, *COPB2*, *ARCN1*, *COPG1*, *COPG2*, *COPE*, *COPZ1*, *COPZ2, LDLR, ABCA1, SCARB1*) or non-targeting siRNAs (please see Major Resources Table for details). Briefly, the siRNAs were diluted in water while in parallel, RNAiMax (cat. 13778150, Thermo Fisher Scientific) was diluted in DMEM (0.075 µL of RNAiMax for each 10 µL of DMEM). One volume of siRNA mix and two volumes of diluted RNAimax were mixed and incubated for 1 hour at room temperature; thereafter the mix was distributed to the plates. At the same time, Huh-7 cells were detached by incubation with 0.5 mg/mL trypsin and 0.2 mg/mL EDTA (cat. T4174, Sigma-Aldrich) for 15 minutes at 37°C, resuspended in DMEM supplemented with FBS and without antibiotics, counted using Vi-Cell XR cell viability analyzer (cat. 731196, Beckman Coulter) and seeded. The final concentration for each siRNA after the addition of the cell suspension was 10 nM. Thereafter, the cells were cultured for 72 hours in a humidified incubator (37°C and 5% $CO_2$), unless otherwise specified. Knockdown efficiency was determined by qRT-PCR.

**Generation of Huh-7 cells overexpressing GFP-SR-BI**

The plasmid containing the coding sequence of *SCARB1* (NM_005505) carrying an N-terminal GFP tag  was purchased GeneCopoeia ( cat. EX-G0782-M29, Rockville, USA),. On Day 1, Huh-7 cells were seeded into 6-well plates using normal growth medium. On Day 2, the medium of the cells was changed to normal growth medium without antibiotics (DMEM +10%FBS, - P/S) and the cells were transfected with 3 µg of an empty plasmid (cat. EX-NEG-M02, GeneCopoeia) or the plasmid encoding for GFP-SR-BI using 6 µL of Lipofectamine 2,000 µL medium /well (cat. 11668019, Invitrogen, Waltham, USA) according to the manufacturer's instructions. 6 hours post transfection, the medium was removed and fresh normal growth medium without antibiotics was added. Finally, 24 hours after transfection the culture medium was replaced with selection medium [(DMEM +10% FBS +1% P/S) supplemented with 750 µg/mL G418 (cat. 10131-027, Gibco, Waltham, USA)]. Cells were selected over 4 passages and maintained under low confluence before being sorted using a BD FACS ARIA III (BD-Biosciences. Allschwil, Switzerland). Single sorted GFP-positive$^+$ cells were then cultured in 96-well plates to obtain single-cell derived colonies. Cells transfected with the empty plasmid served as negative control (EV).

**Real-Time Quantitative Reverse Transcription PCR (qRT-PCR)**

Huh-7 cells were harvested using Tri reagent® (cat. T9424, Sigma-Aldrich,) and the extracted RNA was treated with DNAse I (cat. 04 716 728 001, Roche, Basel, Switzerland) and cDNAs were generated using the RevertAid First Strand Synthesis kit (cat. K1621, Thermo Fisher Scientific) according to manufacturer`s instructions. Quantitative real time PCR reactions were carried out on a Roche Light Cycler 480-II (cat. 05015243001, Roche,) using the LightCycler® 480 SYBR Green I Master (cat. 04887352001, Roche). For each run, the following thermocycle conditions were used: pre-incubation at 95°C for 5 minutes and 45 cycles of denaturation at 95°C for 10 seconds, primer annealing at 60°C for 10 seconds and extension at 72°C for 20 seconds. For each experiment, 3 technical replicates were used for each condition. At the end of each PCR run, the specificity of the PCR products was confirmed by melting temperature (Tm) analysis. The data were analyzed by performing relative quantification based on crossing point (Cp) values for the reference gene (GAPDH) and the gene of interest using the relative standard curve method. For each condition, the ratio of the signal for each gene of interest / signal GAPDH was calculated and the data were normalized to the respective control condition, as described for each experiment. Primer sequences and their respective target genes are described in Supplemental Table S4.

**Uptake of fluorescently labeled HDL**

72 hours after transfection with the indicated siRNAs, the cells were incubated at 37°C with 20 µg/mL of Atto655- / Atto647-HDL or DiI-HDL in DMEM supplemented with 0.2% BSA in the presence (unspecific uptake) or absence (total uptake) of 100 times excess of the respective unlabeled lipoprotein. After 3 hours ( both Atto655- / Atto647-HDL and DiI-HDL), the cells were extensively washed with PBS and detached by incubation with Accutase® (cat. A6964, Sigma-Aldrich) for 5 minutes at 37°C. After washing with PBS and in order to exclude signal from dead cells, the samples were incubated with Propidium Iodide (PI) (cat. 81845, Fluka, Buchs, Switzerland) and FITC-Annexin V (cat. 640945, Biolegend, San Diego, USA) at final concentrations of 1 µg/mL and 2 µg/mL respectively. Sample acquisition was carried out on a BD LSR II Fortessa (BD-Biosciences,) and using BD FACSDIVA™ software. During acquisition, signal from cell debris was excluded by applying the forward size scatter versus the side size scatter gating (FSC-A / SSC-A). Thereafter, the doublets were also excluded through gating (FSC-A / FSC-H). To exclude signal originating from the dead cells, the double negative cells for both FITC-Annexin V and PI were selected by gating (FITC-A / PI-A). To account for the spectral overlap of the different fluorophores used, unstained and single stained controls were acquired and analyzed using the compensation function of the FACSDIVA™ software. In all experiments, approximately $10^4$ events per condition recorded at the final gate containing the population of alive cells were used for analysis. Data analysis was carried out using FlowJo version 10 (FlowJo LLC, Ashland, USA). The Median Fluorescence Intensity (MFI) of each population was used for comparison between the different conditions and the specific uptake was calculated as the difference between total uptake and unspecific uptake for each condition.

**Flow cytometry of cell surface SR-BI and overall emitted GFP signal**

SR-BI and the overall emitted GFP signal were determined by flow cytometry using alive Huh-7 cells overexpressing either GFP-SR-BI or an empty vector. 72 hours after transfection, the medium was aspirated and the cells were washed twice with PBS and detached using Accutase® (cat. A6964, Sigma-Aldrich) for 5 minutes at 37°C. The cells were then washed in ice cold PBS and incubated in PBS containing 0.5% BSA and 2% FBS on ice. After 45 minutes, the cells were incubated with anti-SR-BI antibody in FACS buffer (PBS containing 0.5% BSA and 0.05% $NaN_3$) on ice. After 1 hour, the cells were incubated with a donkey anti-rabbit secondary antibody in FACS buffer for 1 hour on ice in the dark. Finally, after washing twice with FACS buffer and in order to exclude signal originating from dead cells, the cells were incubated with either a combination of PI and FITC-Annexin V (Huh-7 cells transfected with siRNAs) or with PI only (Huh-7 cells overexpressing the empty vector or the GFP-SR-BI construct), as described above. Cells incubated only with the secondary antibody were used as the negative control. During acquisition of cells for measurement of the cell surface signal, the emitted GFP signal was also recorded. Sample acquisition, compensation for spectral overlap, and data analysis were carried out as described above. The gating strategy for the wild type Huh-7 cells was as described above; for the Huh-7 cells overexpressing the empty vector or the GFP-SR-BI construct, the signal from dead cells was eliminated by excluding the PI positive cell population. Sources and concentrations of both primary and secondary antibodies used are summarized in the Major Resources Table.

**Western Blotting**

Huh-7 cells were lysed using radioimmunoprecipitation (RIPA) buffer (25 mM Tris pH 7.6, 150 mM sodium chloride, 24.1 mM (1% w/v)sodium deoxycholate, 0.1% NP-40 and 0.1% SDS) supplemented with COMPLETE$^{TM}$ protease inhibitors (cat. 11836153001, Roche). Protein of the lysates was measured by using BC assay protein quantification (cat. UP40840A, Interchim, Montluçon, France). Equal amounts of total protein were separated by SDS-PAGE followed by electrophoretic transfer to Amersham Hybond 0.45 polyvinylidene fluoride (PVDF) membrane (GE Healthcare Illinois, USA). Antibodies were dissolved in PBS containing 0.01% Tween®-20 (cat. P1379, Sigma-Aldrich) (PBST) with 5% skim milk. The membranes were incubated with primary antibodies overnight at 4°C. After three washes with PBST, the membranes were incubated with secondary antibodies for 2 hours at room temperature. After three additional washes with PBST, the membranes were developed using Thermofisher Scientific SuperSignal™ West Pico PLUS Chemiluminescent Substrate (cat. 34577, Thermo Fisher Scientific) and a Fusion FX imaging system (Vilber Lourmat, Collégien, France). The intensity of the bands was recorded by densitometry using ImageJ[55]. Sources and concentrations of both primary and secondary antibodies used are summarized in the Major Resources Table.

**Cell Surface Biotinylation (CSB)**

48 hours after transfection with the indicated siRNAs, Huh-7 cells were treated with DMEM supplemented with 0.5% FBS, 1% P/S and 10 µM T0901317. 24 hours later, the cells were washed with ice cold PBS, cooled down for 5 minutes on ice and then incubated with ice cold PBS containing EZ-Link sulfo-NHS-S-S-Biotin (cat. 21441, Thermo Fisher Scientific) at a final concentration of 85 µg/mL with mild shaking. After 1 hour, the cells were washed once with ice-cold Tris (50 mM, pH 7.4) and twice with PBS containing 0.1 mM $CaCl_2$ and 1

mM $MgCl_2$. The cells were then harvested into PBS by scraping, pelleted by centrifugation at 700 g, for 5 minutes at 4°C and lysed by using RIPA buffer (total cell lysate). 100 µg of lysates from each sample were incubated with 50 µL of BSA-blocked streptavidin beads suspension (cat. 17-5113-01, GE Healthcare) overnight at 4 °C. The beads were then thoroughly washed 5 times with RIPA buffer. For the dissociation of the biotinylated surface proteins, the beads were incubated with 6x SDS loading buffer (350 mM Tris pH=6.8, 30% glycerol, 10% w/v SDS, 9.3% w/v DTT, 0.012% w/v Bromophenol blue) for 30 minutes at 40°C. The samples were then centrifuged and the supernatants containing the dissociated surface proteins were collected and separated by SDS-PAGE and immunoblotted as described above.

**Enzymatic deglycosylation**

Huh-7 cells transfected with the indicated siRNAs were harvested into PBS by scraping, pelleted by centrifugation and lysed with RIPA buffer 72 hours after transfection. 20 µg of lysates were incubated with PNGase F, neuraminidase and/or O-glycosidase (New England Biolabs, Ipswich, USA cat. P0704, cat. P0720, and cat. P0733, respectively) for 9 hours at 37°C, according to the manufacturer's instructions. Equal amounts from each sample were used for SDS-PAGE and immunoblotting as described above.

**Confocal Microscopy**

Wild type Huh-7 cells or Huh-7 cells overexpressing or GFP-SR-BI were seeded in 24 well plates containing glass coverslips and transfected with the indicated siRNA. After 72 hours, the cells were fixed in PBS containing 4% paraformaldehyde (PFA) for 20 minutes at room temperature. The cells were then washed 3 times with PBS and incubated with PBS containing 0.1 % saponin (cat. 84510, Fluka, Buchs, Switzerland) for 10 minutes at room temperature. After three washes with PBS, the cells were incubated in PBS containing 5% normal donkey serum (cat. 566460, Sigma- Aldrich) and 1% BSA for 1 hour. The cells were then incubated with primary antibodies diluted in PBS containing 5% normal donkey serum and 1% BSA overnight at 4°C. After five washes with PBS, the cells were incubated with secondary antibodies. in PBS containing 5% normal donkey serum and 1% BSA for 45 minutes at room temperature. After five additional washes with PBS, the cells were mounted in Prolong™ Gold Antifade reagent with DAPI (cat. P36935, Invitrogen) and were imaged using a SP8 confocal microscopy (Leica, Weltzar, Germany). For the lysotracker assay, the cells were incubated for 30 minutes with 10 nM Lysotracker® Red DND-99 (cat. L7528, Molecular Probes, Oregon, USA) diluted in DMEM without FBS before being processed as above with the exception of no permeabilization. Sources and concentrations of both primary and secondary antibodies used are summarized in the Major Resources Table.

**Quantification of apoA-I secreted by Huh-7 cells**

For quantification of apoA-I secretion by ELISA, Huh-7 cells transfected with siRNAs were cultured for 48 hours, detached using trypsin-EDTA, counted, and re-seeded into 6-well plates (2 wells per condition), to account for any differences in cell number between the different conditions until that point. 3 hours after reseeding, the media of the cells were replaced with DMEM (cat. D1145, Sigma-Aldrich) supplemented with 1% P/S but free of phenol red and FBS (1 mL/6-well). After an additional 48 hours, both the media and the cells of each condition were collected. To remove any dead cells, the media were centrifuged twice (500 g, 5 minutes each round). The infranatants (approximately 200 µL after each centrifugation step per condition), which contained the pelleted floating dead cells were pooled with the originally collected cells for counting and normalization of the apoA-I concentration in the media. The remaining media volume ( at least 1.3 mL for each condition) were collected and used to determine the concentrations of apoA-I in the cell culture media using a commercially available ELISA kit (cat. 3710-1HP Mabtech Nacka Strand, Sweden), according to the manufacturers's instructions.

**Cholesterol efflux**

Huh-7 cells were seeded into 24-well plates and transfected with siRNAs, as described above. 48 hours post transfection, the cells were incubated overnight with 500 µL of DMEM containing 1 µCi/mL [3H]-cholesterol (cat. NET139001MC, PerkinElmer, Waltham, USA), FBS (ratio 1:40), 10 µM T0901317 (cat. T2320, Sigma-Aldrich) and 5 µg/mL Sandoz 58-035 (cat. S9318, Sigma-Aldrich). On the next day, the medium was removed and the cells were washed with DMEM supplemented with 1% BSA before being incubated with 500 µL of DMEM supplemented with 0.2% BSA, 10 µM T0901317 and 5 µg/mL Sandoz 58-035. After 3 hours the medium was removed, the cells were washed with DMEM +1% BSA and incubated with 500 µL of DMEM containing 5 µg/ml Sandoz 58-035 without (no acceptor) or with 50 µg/mL of HDL or 25 µg/mL of apoA-I as acceptors. After 4 hours, 450 µL of medium from each well were collected and the cells were washed extensively with PBS before lysis with 0.1 M NaOH. Assay medium and cell lysates were measured using a 2250-CA Tri-Carb liquid scintillation analyzer (Packard, USA). The percent cholesterol efflux was calculated as (cpm in medium/[cpm in the cell + medium]) × 100. For each condition, cholesterol efflux obtained in the absence of any acceptor was subtracted from the cholesterol efflux obtained in the presence of either apoA-I or HDL.

**Gas chromatography – mass spectrometry (GC-MS) of cellular sterols**

48 hours after transfection of Huh7 cells with siRNAs, the medium was replaced with DMEM supplemented with 0.5% FBS, 1% P/S, 50 µg cholesterol /mL of HDL in the absence (HDL uptake) or in the presence of 10 µM T0901317 and 5 µg/mL Santoz 58-035 (HDL efflux). After 24 hours, the cells were collected and pelleted. Sterols were extracted from dried cells with chloroform overnight. Cholesterol, its precursors (lanosterol, dihydrolanosterol, desmosterol, and lathosterol) as well as phytosterols (campesterol, stigmasterol, and sitosterol) were separated and measured after trimethylsilylation by gas chromatography-mass spectrometry- selected ion monitoring (GC) as described in detail previously[56,57]. The amounts of each sterol in the cells was normalized to the actual mass of dry cell pellet.

**Thin layer chromatography (TLC) of unesterified and esterified cholesterol in cells**

48 hours after transfection of Huh7 cells with siRNAs, the medium was aspirated and replaced with DMEM (cat. D-5796, Sigma-Aldrich) supplemented with 0.5% FBS and the indicated combinations of  DMSO, 50 µg cholesterol/mL HDL, 10µM LXR agonist and 5 µg/mL ACAT inhibitor for 24 hours. After 24 hours, the cells were harvested and counted with a Vi-CELL XR Cell Viability Counter, (Beckman Coulter,). Approximately $10^6$ cells were resuspended in 1 mL of PBS spiked with 0.1 µCi/mL [14C]-Cholesterol (cat. NEC018250UC,

specific activity: 50.8 mCi/mmol, PerkinElmer) -serving as an internal standard- and transferred into a glass tube. 50 µL of cell suspension was mixed with 50 µL of 1% (w/v) Triton X-100 in deionized water for bicinchoninic acid protein determination (cat. UP95424A and UP95425A, Interchim). The remaining cell suspension was mixed with 3 mL of chloroform:methanol (2:1) solution and was shaken for 20 minutes at room temperature. Thereafter the samples were centrifuged for 5 minutes at 500 g for phase separation. The upper phase and interphase were carefully aspirated, and the lower phase containing the lipid fraction was dried under a nitrogen flux at 30℃. The glass tubes containing the dried lipids were cooled for approximately 30 minutes at 4°C and then resuspended in 300 µL of ice-cold chloroform. A 25µL aliquot was used for assessing radioactivity by liquid scintillation counting (Tri-Carb 2250 CA, Canberra Packard, Schwadorf, Austria). For the analysis of the cholesterol and cholesteryl ester, samples were loaded on HPTLC Silica gel 60 plates with a concentrating zone (cat.1.05626.0001, Merck Rahway, USA) using an automated Camag TLC sampler ATS4 (Muttenz, Switzerland) and separated by one-dimensional thin layer chromatography (TLC). Cholesterol and cholesteryl ester were resolved in a mobile phase containing 18.7 mL n-hexane, 5.5 mL n-heptane, 5.5 mL diethyl ether, and 0.3 mL of acetic acid. Staining was performed with 10% (v/v) orthophosphoric acid and 3% (w/v) copper acetate. Finally, lipids were charred at 170-180℃ for 25-30 minutes for band visualization. Bands were scanned at 366 nm in a Camag TLC Scanner 3 (Muttenz, Switzerland) and quantified from a cubic function generated from a serial dilution of cholesterol (cat. C3045, Sigma-Aldrich) and cholesteryl oleate (cat. C9253, Sigma-Aldrich). The absolute values were then corrected for the [14C]-Cholesterol signal and normalized for the protein concentration.

**Human genetic studies**

The GWAS summary statistics for variants and phenotypes were downloaded from http://csg.sph.umich.edu/willer/public/glgc-lipids2021/results/trans_ancestry/. The data included meta-analysis of associations of 51,735,689 detected and imputed SNPs from approx. 1.65 million subjects from five different ethnicities as described by the Global Lipids Genetics Consortium[17]. Variants were assigned to each gene using the UCSC Genome Browser Variant Annotation Integrator (https://genome.ucsc.edu/cgi-bin/hgVai), build GRCh37/h19. Only variants between the transcription start and end were considered. Supplementary table S5 shows the transcription start and end positions for the selected genes. METAL p values were corrected using the Bonferroni correction method based on the total number of SNPs in the whole dataset.

Associations between UK biobank exome variants in the selected genes were downloaded from https://genebass.org/ [18]. The association p value was corrected using the Bonferoni correction based on the number of 8,553 variants in the selected COPI genes. Data analysis visualization was done in R (R-project.org) using the packages ggplot,data.table and dplyr.

Five patients with the COPA syndrome due to heterozygosity for the rare mutations in the *COPA* gene presented in figure 6B as well as the patients with combined immunodeficiency due to homozygosity for the rare p.K652E mutation in the *COPG1* gene and there heterozygous parents presented in figure 6C were previously described[8,9]. Data on demographics and lipids were provided by Drs. Anthony Shum (San Francisco), Raif Geha and Janet Chou (both Children's Hospital in Boston, USA). Plasma lipids were analyzed by the use of enzymatic photometric assays from Abbott (Architect Chemistry Analyzer) and Sigma in San Francisco and Boston, respectively. Because of their different geographic and ethnic origin, we compared HDL-C levels in COPA and COPG1 variant carriers with those in different populations, namely NHANES from the United States[19] for the COPA variants and population studies in adults and children from the United Arab Emirates for the COPG1 variants[20,21].

**Animal studies**

Heterozygous $Copa^{wt/E241K}$ and homozygous $Copg1^{K652E/K652E}$ mice have been previously described[9,22] and were maintained in the specific pathogen free facilities at UCSF and Boston Children's Hospital, respectively. Blood for plasma preparations were collected in the US laboratories. Frozen plasmas were shipped to Groningen for the profiling of plasma lipoproteins by fast protein liquid chromatography (FPLC) as described previously[58]. Pools of equal plasma volumes from 3 male or 3 female $Copa^{wt/E241K}$ and 5 male or 5 female $Copg1^{K652E/K652E}$ mice were passed through a SuperoseTM 6 Increase 10/300 GL column (GE Healthcare Hoevelaken, Netherlands) at a flow rate of 0.31 mL/minute for lipoprotein fraction separation. Chromatographic profiles of pooled wildtype (C57Bl/6) mice served as a reference standard. Data analysis was performed using GraphPad Prism 10.

**Statistics**

The RNAi screening assay feature data were analyzed as follows: data were first normalized by the median value of each batch, microscope, plate and well and were finally expressed as robust Z-score[59] normalized values. The Redundant siRNA Activity (RSA) analysis was performed for each assay feature on the normalized data to rank the genes and detect the top hits defined here as the genes with an RSA p- value of less than $10^{-3}$. This p- value cutoff was dictated by our ability to verify the results in vitro. Transfection efficiency and the dynamic range of the screening assay were determined by calculating the Z'-factor between positive and negative transfection and assay controls as published before[59].

All experimental data were analyzed using GraphPad Prism version 10. With the exception of the data of figure 4H, all results with $n<5$ were analyzed with non-parametric tests, namely Mann-Whitney test for one to one comparisons or Kruskal-Wallis test coupled with Dunn's test for multiple comparisons when comparing several conditions. Results with $n\geq5$ were first evaluated for normal distribution of all depicted conditions of one experiment using Shapiro-Wilk normality test. If all examined conditions were normally distributed, the data were analyzed with parametric tests, namely unpaired t-test and 1-way ANOVA coupled with either Dunnett's or Bonferroni's test for multiple comparisons when comparing several conditions, as indicated in the respective figure legends. In case of not normal distribution even for a single condition within one experiment, the data were analyzed with non-parametric tests, as described above. For graphs in which all depicted conditions were compared to the respective control, only p values lower than the threshold limit mentioned in each legend are depicted. Otherwise, if different conditions were compared with each other, all p values are depicted regardless of statistical significance. All p values were rounded to the third significant decimal digit. The numbers of experiments, the used statistical tests, and, if applicable, adjustments for multiple testing are described in the legends of the respective figures and tables.

**Ethics**

The use of clinical data and samples from patients with the COPA syndrome or the COPG1 syndrome for this study was approved by the Institutional Review Boards (IRB) for the protection of human subjects of the University of California in San Francisco (UCSF, IRB protocol 10-02467) and Boston Children’s Hospital (IRB protocol *04-09-113R*). All participants provided written informed consent.

The sampling of blood and livers from the mutant COPA and COPG1 mice were performed as approved by the Institutional Animal Care and Use Committees of UCSF in San Francisco (Mouse protocol number and approval number 202539) and Boston Children’s Hospital (Mouse protocol number and approval number 00001617), respectively.

### Supplemental figures

A

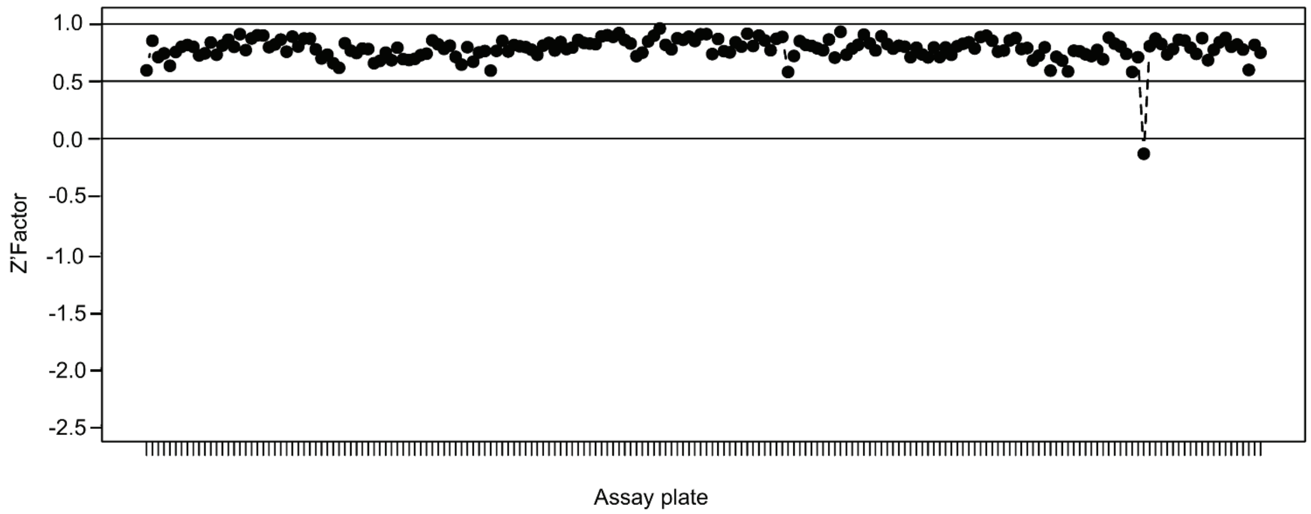


**Supplemental Figure S1. Quality control (QC) of the screening.** This figure depicts the Z'-factors for median cytoplasm intensity in the CY5 channel for negative control wells that did not receive any fl-HDL. The Z'-factor was calculated in comparison to wells that received a non-targeting control siRNA and were subsequently incubated with fl-HDL. Z'-factors between 0.5 and 1 are considered excellent.

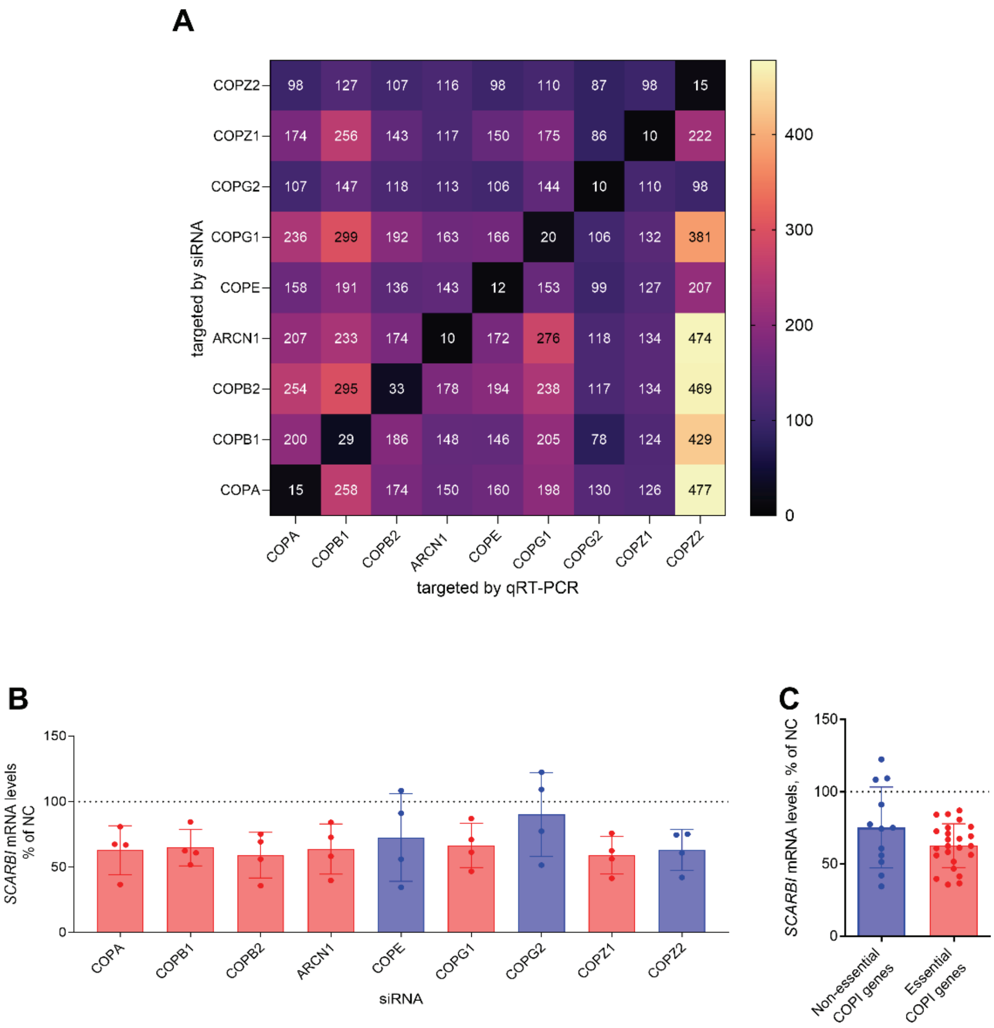


**Supplemental Figure S2. Effects of silencing the different COPI subunits on mRNA levels of COPI genes (A) and *SCARB1 (*B,C*).*** Huh-7 cells transfected with the indicated siRNAs were collected 72 hours post transfection and used for the quantification of the indicated mRNAs by using qRT-PCR. In all cases, the data were normalized to the non-coding (NC) control and are shown either as a heatmap with each numerical value representing the mean of 3 independent experiments (**A**) or as means ± SD of 4 independent experiments (**B**). In **A** the x axis describes the COPI genes targeted by qRT-PCR, the y axis the COPI genes targeted by RNAi and the numbers indicate the means of the 3 independent experiments. **C** compares the summarized data on the non-dispensable COPI genes (red bars) with the data on the dispensable or paralogous COPI genes (blue bars). Statistical analysis using either Kruskal-Wallis test with Dunn's test for multiple comparison between the NC and each targeting siRNA (**B**) or two-tailed unpaired t-test (**C**) did not reveal any statistical significant difference.

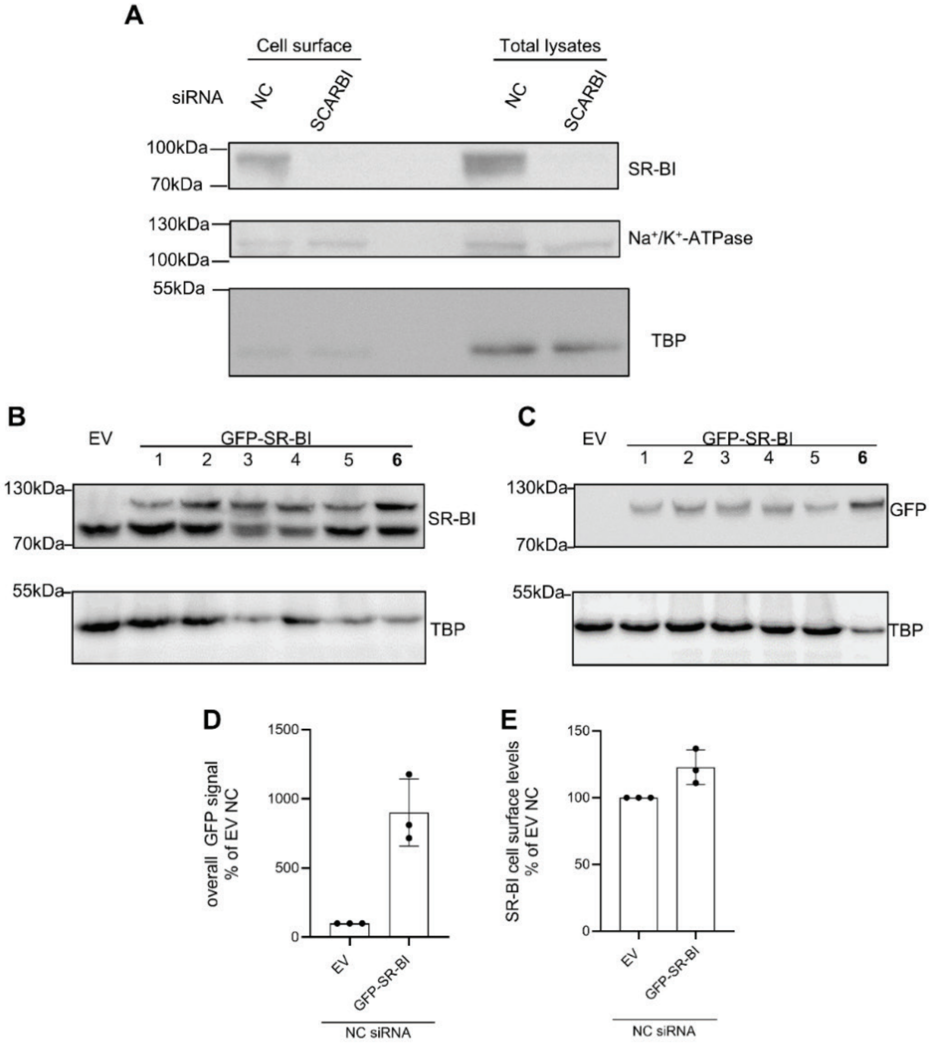


**Supplemental Figure S3. Evaluation of SR-BI's total and cell surface abundance in Huh-7 cells without or with stable overexpression of GFP-SR-BI.** Wild-type Huh-7 cells (**A**) or Huh-7 cells stably overexpressing GFP-SR-BI (**D**,**E**) were transfected with the indicated siRNAs (**A**,**D**,**E**) .**A.** After 72 hours, the cells were used for cell surface biotinylation experiments. The blot was probed with the indicated antibodies, with the anti-TBP serving as a control for the intracellular protein expression and $Na^{+}/K^{+}$ ATPase as a control for surface protein expression. For details see methods. **B-E** Huh-7 cells stably overexpressing an empty vector (EV) or a plasmid encoding for GFP-SR-BI were either lysed for Western Blotting and probed with antibodies against either SR-BI (**B**) or GFP (**C**), or analysed by flow cytometry of the GFP signal (**D**) or the anti-SR-BI immunoreactivity on the cell surface (**E**). The numbers in **B** and **C** indicate individual single-cell derived clones of Huh-7 cells overexpressing GFP-SR-BI. Cells of clone "**6**" showed highest overexpression and were propagated for further experiments**.** The Western Blotting experiments in **A-C** were carried out once. In both **D** and **E** the data were normalized to the cells overexpressing the empty vector (EV) and transfected with non-coding (NC) siRNA and are shown as means ± SD of 3 independent experiments.

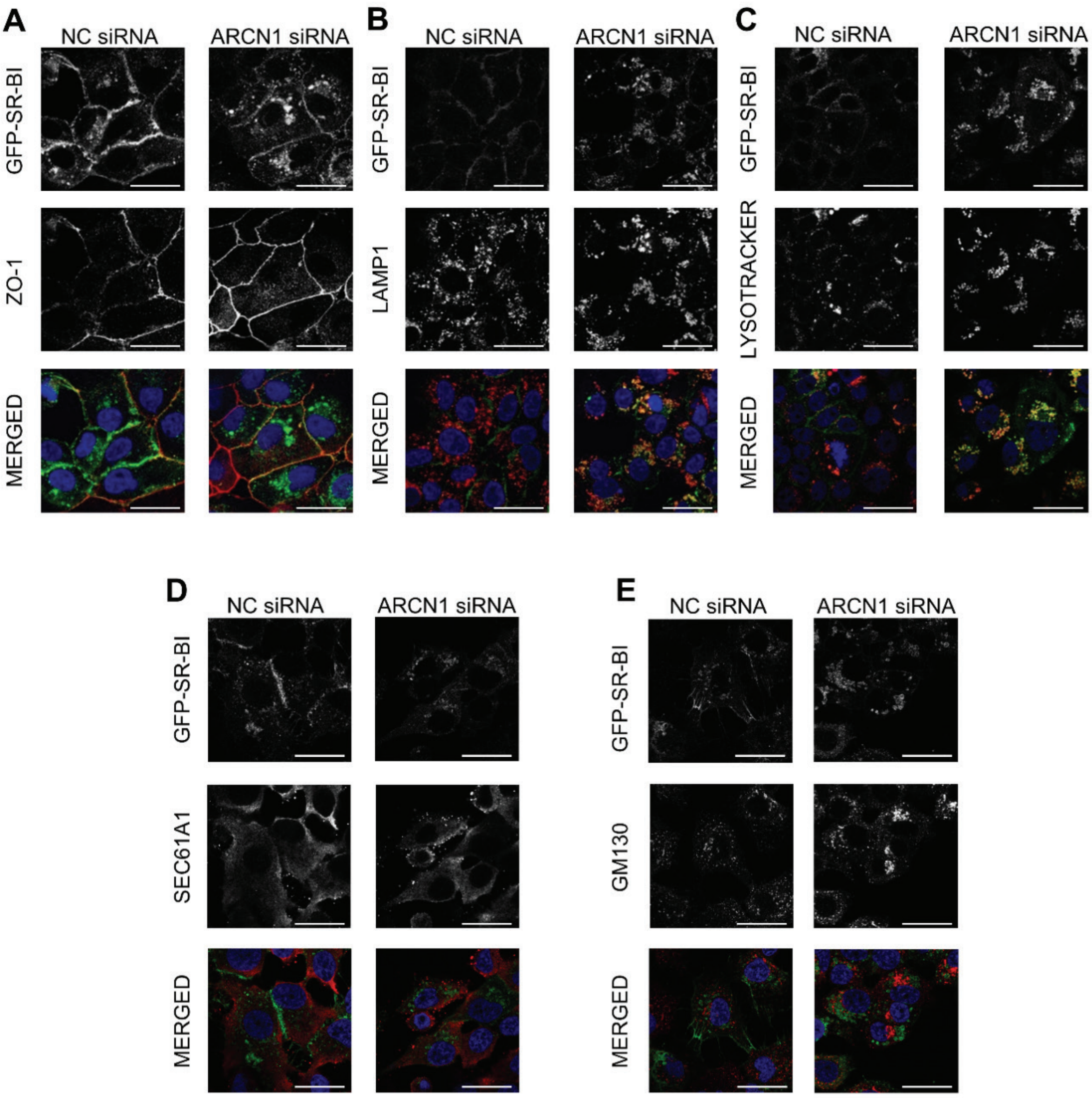


**Supplemental Figure S4. Loss of ARCN1 alters the intracellular localization of SR-BI in Huh-7 cells**. Confocal micrographs of GFP-SR-BI and ZO-1 (**A**), LAMP1 (**B**), LYSOTRACKER (**C**), SEC61A1 (**D**) and GM130 (**E**) of Huh-7 cells overexpressing GFP-SR-BI 72 hours after transfection with non-coding siRNA (NC) or anti-ARCN1 siRNA. The micrographs of the merged signal in **A** and **B** are the ones presented in Figures 2H and 2I, respectively. The data shown represent two independent experiments. The scale bar for all micrographs is 25 μm.

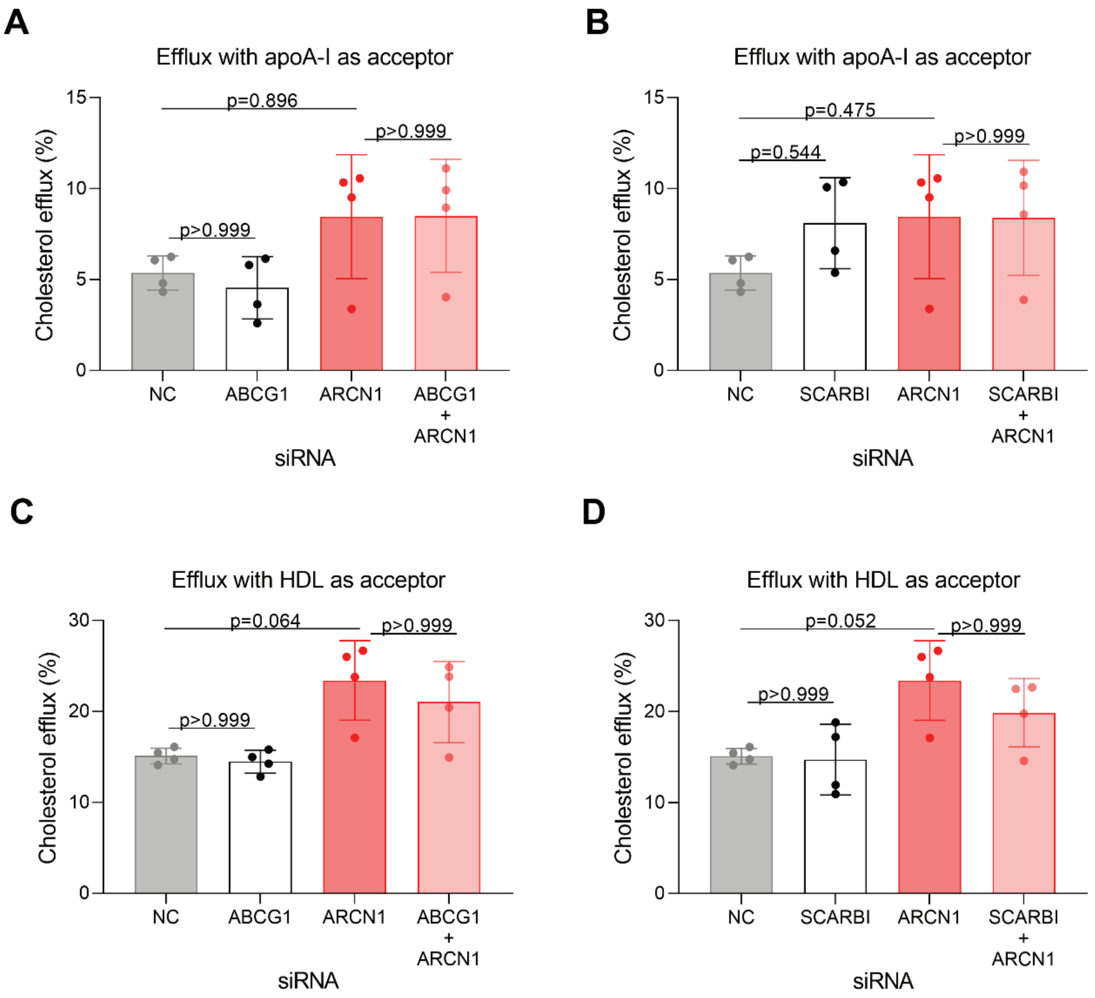


**Supplemental Figure S5. ABCG1 and SR-BI are not involved in the increased cholesterol efflux observed upon silencing *ARCN1*.** The efflux of [$^3$H]-cholesterol was measured 72 hours after transfection of Huh-7 cells with the indicated siRNAs by using lipid-free apoA-I (**A**,**B**) or HDL (**C**,**D**) as the acceptors. In all cases, the data are shown as means ± SD of 4 independent experiments. Statistical analysis was performed using Kruskal-Wallis test coupled with Dunn's test for multiple comparisons between the indicated conditions.

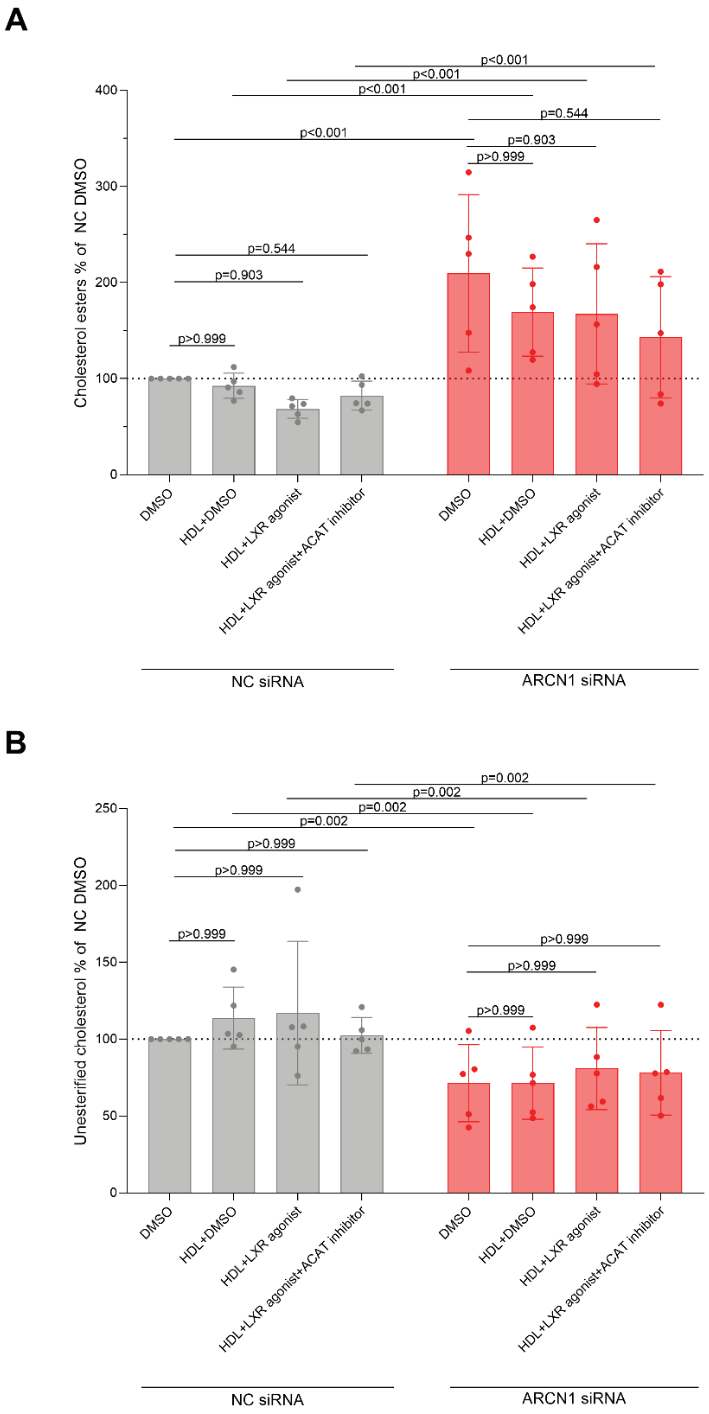


**Supplemental Figure S6. Silencing *ARCN1* alters both the unesterified cholesterol and the cholesterol ester content of Huh-7 cells.** 48 hours after transfection with the indicated siRNAs, the cells were treated with DMEM supplemented with 0.5% FBS and the indicated combinations of DMSO, 50 µg cholesterol/mL HDL, 10µM LXR agonist and 5 µg/mL ACAT inhibitor for 24 hours. Thereafter, the cells were collected for separation of unesterified and esterified cholesterol by TLC. The data were normalized to NC DMSO and are shown as means ± SD of 5 independent experiments. Statistical analysis was performed using 2-way ANOVA coupled with Bonferroni's test for multiple comparisons and only the indicated conditions.

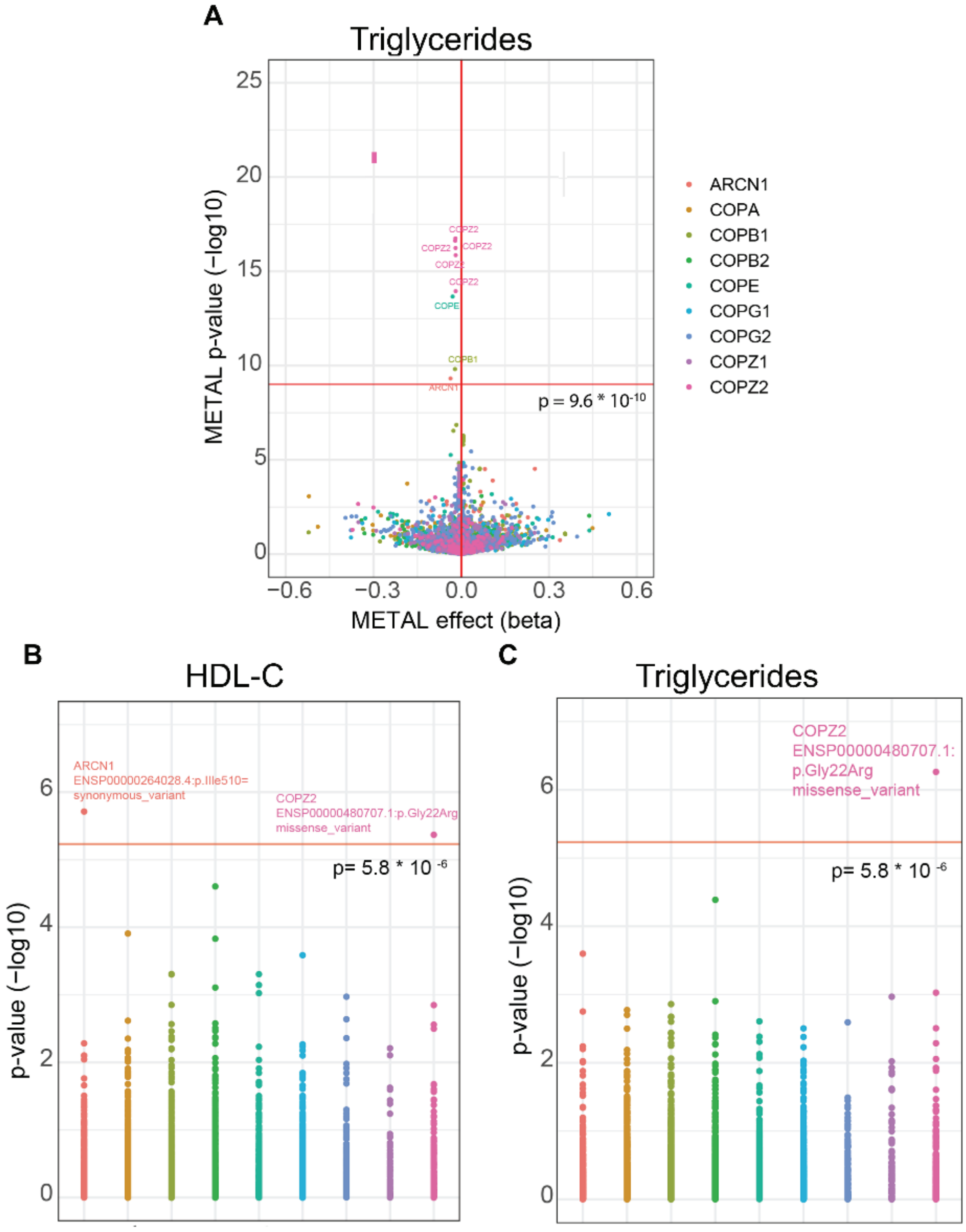


**Supplemental Figure S7. Associations of SNPs (A) or rare exome variants (B,C) of COPI genes with triglycerides (A,C) and HDL-C (B) in the general population.** Volcano plot **(A)** depicting the association of 5,618 SNPs of the nine COPI genes with triglycerides in 1.65 million individuals aggregated by the Global Lipid Genetics Consortium[17]. The x-axis depicts the effect size (beta) and the y-axis the p-value. The dashed horizontal line marks the threshold of statistical significance after Bonferroni correction for multiple testing on GWAS level ($p = 9.60*10^{-10}$). **B** and **C** show associations of rare exome variants of COPI genes with plasma levels of HDL-C and triglycerides, respectively, in > 394,000 subjects of the UK biobank database[18]. The y-axis indicates the p values. The dashed line indicates the threshold for statistical significance after Bonferroni correction for multiple testing of 8,553 variants ($p=5.8*10^{-6}$).

### 3. Supplemental Tables

**Supplemental table S1: Complete screening dataset for the five best performing assay features (provided as Excel file).** The assay features values are normalized as reported in the methods section. Non self-explanatory column names are as follows: siRNA.ID: siRNA molecule ID as according to manufacturer; Gene_ID: NCBI Gene-ID on 2016.03.14; Batch: experimental batch number; RefSeq.Accession.Number: NCBI RefSeq ID for the targeted transcript; Gene.Symbol.20160314: NCBI Gene Symbol on 2016.03.14; FL.LABEL: describes the fluorescent labels added to each well; RNASeq_Huh-72..Signal.: normalized gene-level expression as measured by RNA sequencing; RNASeq_Huh-72..Present.: dichotomic gene-level expression label (threshold 7.5). For each assay feature, RSA p values are given for both directionalities of the RSA analysis (inhibition and enhancement of HDL uptake). Empty wells, control wells, wells without an associated Gene.ID as well as wells that did not meet the lower ranking threshold for the RSA analysis are not shown, as no RSA p-value was calculated for those.

**Supplemental table 2. Effects of *ARCN1* knockdown on cellular levels of sterols in Huh-7 cells.**

| classification | sterol | HDL uptake | | | Cholesterol efflux | | |
|---|---|---|---|---|---|---|---|
| | | NC (ng/mg cell pellet) | ARCN1 (ng/mg cell pellet) | P | NC (ng/mg cell pellet) | ARCN1 (ng/mg cell pellet) | P |
| cholesterol precursors | lathosterol | 1,039±145 | 1,097±67 | 0.700 | 1517 ±371 | 1360±56 | 0.700 |
| | lanosterol | 48.133±15.610 | 70.867±22.497 | 0.400 | 122.000±15.621 | 180.333±14.295 | 0.100 |
| | desmosterol | 75.100±10.912 | 88.100±3.404 | 0.300 | 81.767±5.401 | 100.633±11.018 | 0.100 |
| | dihydrolanosterol | 2.053±0.541 | 4.663±1.644 | 0.100 | 3.973±0.415 | 7.357±0.855 | 0.100 |
| | cholesterol | 29,533±7,853 | 22,700±2,022 | 0.400 | 23,267±3,350 | 17,767±635 | 0.100 |
| phytosterols | stigmasterol | 1.500±0.255 | 1.860±0.131 | 0.200 | 1.907±0.451 | 2.120±0.139 | >0.999 |
| | sitosterol | 14.053±5.102 | 6.130±1.881 | 0.200 | 10.890±1.287 | 7.513±0.480 | 0.200 |
| | campesterol | 10.900±0.424 | 7.070±0.270 | 0.200 | 6.397±0.311 | 3.787±0.868 | 0.100 |

The indicated sterols were measured by gas chromatography-mass spectrometry in lipid extracts of pellets from Huh-7 cells which were transfected with siRNAs targeting ARCN1 or non targeting siRNAs (NC) and incubated with HDL in the absence ("HDL uptake") or presence ("cholesterol efflux") of an LXR agonist and ACAT inhibitor to reflect the conditions used for the recording of HDL uptake and cholesterol efflux, respectively**.**

The data are shown as mean ± SD of 3 independent experiments. The p values presented are calculated with Mann-Whitney test (two-tailed) and are not adjusted for multiple testing;

**Supplemental table S3. PCR primer sequences for all primer pairs**

| Primer name | 5'-3' Sequence |
|---|---|
| GAPDH-F | CCCATGTTCGTCATGGGTGT |
| GAPDH-R | TGGTCATGAGTCCTTCCACGATA |
| COPA-F | CTAAGCGGACTGGGGTTCAG |
| COPA-R | AACTTGGAACCACTCCGCAA |
| COPB1-F | TCTTTGTTGCTGCCTCCCTT |
| COPB1-R | GCCATGAGCAACATAGCCTC |
| COPB2-F | GTCAGTGGATCGGTGGGTTT |
| COPB2-R | ATCAAGTCGCAGAGGCATGG |
| ARCN1-F | TATCACCATCCCACTCCCGT |
| ARCN1-R | TAAACTCCAGGCTGCCACTC |
| COPE-F | GAGAGAGACGTGGAGAGGGA |
| COPE-R | GGGCGAGGTAGTCAGCAAA |
| COPG1-F | GAGTGCTCTGGCGAAGTTTG |
| COPG1-R | TGGACACAGTCAGACCATTTAGG |
| COPG2-F | CCGAATTGCCAGTCGCTTAC |
| COPG2-R | AGCAGGTGCCAACTCTCTTG |
| COPZ1-F | GCTGCAGTCAGCCAAAGAAC |
| COPZ1-R | CCTGAGAGCATCGATTGGGG |
| COPZ2-F | GTCTGTTCTCACCTGCCTGTT |
| COPZ2-R | CCGCCATCCACAATCTCGT |
| ABCA1-F | GCACTGAGGAAGATGCTGAAA |
| ABCA1-R | AGTTCCTGGAAGGTCTTGTTCAC |
| SCARB1-F | CTGTGGGTGAGATCATGTGG |
| SCARB1-R | GCCAGAAGTCAACCTTGCTC |

F: forward primer, R: reverse primer

**Supplemental table S4. Genomic coordinates for the transcription start and end positions of selected COP genes in GRCh37/h19.**

| Gene | chromosome | Transcription start | Transcription End |
|---|---|---|---|
| ARCN1 | 11 | 118443101 | 118473748 |
| COPA | 1 | 160258376 | 160313354 |
| COPB1 | 11 | 14479048 | 14521441 |
| COPB2 | 3 | 139076432 | 139108522 |
| COPE | 19 | 19010319 | 19030212 |
| COPG1 | 3 | 128968452 | 128996616 |
| COPG2 | 7 | 130146078 | 130353598 |
| COPZ1 | 12 | 54718873 | 54745635 |
| COPZ2 | 17 | 46103532 | 46115168 |
